# An Eye on CAUTI Prevention: Bridging the Gap in the Prevention of Catheter-Associated Urinary Tract Infections

**DOI:** 10.1101/2023.01.21.23284862

**Authors:** Karthik Yeruva, Nat West, Warseal Powell

**Affiliations:** Merit Health River Region Hospital, MS; University of South Alabama, AL

**Keywords:** CAUTI, HAI, Infection, Prevention, Foley, Catheter

## Abstract

**Problem:** The United States Healthcare Systems is burdened heavily by healthcare-associated infections (HAI), as they pose significant risks for increased mortality and morbidity. The most common type of HAI is urinary tract infection (UTI), and these infections are preventable (Strassle et al., 2019). According to the CDC (2021), 12-16% of hospitalized adults will at some point require catheterization; and each day an indwelling catheter remains in place increases the patient’s risks of adverse outcomes by 3-7%. CAUTIs have been linked to increased mortality and morbidity rates across the world. The Centers for Disease Control report urinary tract infections (UTI) lead to more than 13,000 deaths each year (Centers for Disease Control [cdc], 2021).

**Objective:** This quality improvement (QI) project will focus on the prevention of hospital-acquired UTIs, specifically those infections related to indwelling devices such as foley catheters. The purpose of this manuscript is to review the current evidence-based literature related to CAUTI prevention, trial an intervention that parallels the literature in a local hospital and evaluate those results.

**Design:** The researcher focused on the problem of CAUTI, analyzed current evidence-based practices related to prevention, developed a plan to execute a high-value improvement tool, and evaluated its effectiveness.

**Setting:** QI project took place in a local community hospital. The focus area was the medical-surgical ICU.

**Participants:** The participants were nursing staff and nursing managers on the medical surgical units, specifically those in positions with unique or direct involvement with insertion, removal, or monitoring of foley catheters. The subjects in this quality improvement project included adult patients hospitalized in the medical surgical intensive care unit (ICU).

**Interventions:** A CAUTI GPS screening tool was used to identify current prevention practices and/or any roadblocks to the prevention of CAUTI within the facility. Then, TAP (targeted assessment for prevention) strategy was implemented, which prompted expedited removal of indwelling catheters and/or the use of external drainage devices where indicated.

**Results:** There was a marked increase in attention to and prompt removal of indwelling foleys throughout the critical care unit. The facility had no hospital-acquired CAUTI during the project period.

**Conclusions:** There must be an impetus to inspire compliance. If healthcare workers adhere to prevention guidelines, CAUTIs are preventable. When leadership team members within hospital systems are enthusiastic about CAUTI prevention, the organization as a whole has increased motivation (Chenoweth et al., 2014).

## Chapter 1: Overview of the Problem of Interest

More than 80% of healthcare-associated UTIs (urinary tract infections) are linked to direct instrumentation of the urinary tract (Institute for Healthcare Improvement, 2021). Catheter-associated urinary tract infections (CAUTI) can be devastating to patients. The ramifications often involve increased discomfort, widespread infection (if not treated), inflated expense, debilitation, and sometimes even death. Roughly 3.2% of hospitalized patients experience HAIs (healthcare-acquired infections). This equates to about 700,000 infections a year, about 1 in every 31 adult patients in the hospital. Treating complications from these infections not only burdens the patients but also can have a huge financial impact on our global and local healthcare systems. The estimated cost is more than $340 million per year (Strassle et al., 2019). Any costs associated with the treatment of CAUTI are considered non-reimbursable (Ferguson, 2018).

Paraplegia, female sex, and stroke are significant risk factors for CAUTI. Despite multitudes of prevention efforts and evidence-based literature being available, there are still significant health ramifications of catheter dwell time and subsequent development of CAUTI throughout hospitals across America. The estimated additional cost to care for patients with CAUTI is $13,793. For every 1000 hospital CAUTIs, there are 36 excess deaths. (Agency for Healthcare Research and Quality [AHRQ], 2017).

The CDC recommends consideration of using external devices in lieu of indwelling foleys when possible; therefore, the proposed project is to implement a CAUTI screening tool that will prompt expedited removal or the use of external drainage devices where indicated. If it is not feasible to avoid using in certain situations (such as obstructive uropathy), then there should be clear parameters set to promote early removal of indwelling foleys (Centers for Disease Control [cdc], 2021). Advanced Practice Registered Nurses (APRNs) can have a strong impact on CAUTI reduction by spearheading quality improvement projects that decrease the use of indwelling catheters. The focus of the manuscript submitted will detail the plans for a quality improvement project aimed at reducing CAUTI in the medical-surgical intensive care unit (ICU) at a local community hospital.

### Background Information

As an incentive to encourage hospital systems to improve their infection reduction programs, the Centers for Medicare & Medicaid Services (CMS) started to place financial penalties on healthcare providers for healthcare-associated infections (HAI). Years later, CMS researchers found that HAI did not actually decrease. What actually happened was that more infections were coded as present on arrival (POA). Numbers as high as 91% of CAUTIs were reported as POA after the 2008 penalty implementation (Calderwood et al., 2018). So there has been essentially minimal impact on reimbursement, but it’s questionable if there has been any real impact on the real reduction of HAI, in general.

One of the most effective ways to address the urgency of healthcare-associated infections is by performing quality improvement projects (Alqarni, 2021). A conversation with the facility’s Chief Nursing Officer (CNO) and Quality Director identified key areas for improvement in the facility, and CAUTI reduction was at the top of the list. The mission of the facility is to provide patient-centered quality care, which includes minimizing adverse outcomes; therefore, the project aligns with the facility’s goals by decreasing the rates of CAUTI among adult patients in the medical-surgical ICU, leading to early detection and treatment to avoid serious complications, such as urosepsis, renal failure, and ultimately the prevention of death. The ultimate goal was to minimize the use of indwelling devices. The reported barrier, per leadership, was nurse compliance.

## Chapter 2: Objectives

### PICO Question

What is the effect of implementing a CAUTI GPS screening tool, as compared to the current screening process, in decreasing the use of indwelling foleys in patients hospitalized in the medical surgical ICU?

### Overarching Aims

Clear outcomes were set, which will include (but were not limited to):

1. increased staff awareness of CAUTI prevention protocols and appropriate indications for catheter usage
2. reduction in the use of indwelling catheters,
3. increased usage of alternative urinary drainage devices
4. decreased number of catheter days of any kind, and
5. a reduction in catheter-associated UTIs.

## Chapter 3: Review of Evidence-Based Literature

Extensive research of the subject area using the University of South Alabama’s Biomedical Library, with search engines such as CINAHL (Cumulative Index to Nursing and Allied Health Literature), PubMed, and Google Scholar, was done using keyword searches, which included: CAUTI, UTI, evidence-based practice, quality improvement, foley catheter, indwelling foley, and infection control. Exclusion criteria were any articles published prior to 2016. There was an abundance of evidence-based literature that supported the use of CAUTI GPS Screening Tools in the clinical setting.

Preventing hospital-acquired infections is a top priority of the CDC. They are weaponizing their efforts with information management tools such as TAP, which can be specifically tailored to focused quality improvement areas, such as CAUTI. The CDC’s National Healthcare Safety Network (NHSN) tracks all TAP data. The 3 parts of the TAP Strategy include: (1) running reports through NHSN to determine which facilities are not meeting standards, (2) using TAP assessment tools within the facilities to help identify gaps in prevention efforts, and (3) utilizing TAP implementation guides to address these deficits. Hospitals nationwide report their information to the NHSN, and each organization is able to run specified HAI reports at any time. The cumulative attributable difference (CAD) is the metric system used by TAP to determine the target number for HAI reduction goals for healthcare systems. Specifically, the CAD represents the number of required preventions to be considered a true reduction rate. Reports can be run to identify the facilities (and units within facilities) that are exceeding national, state, and local level CAUTI rates. This information is publicly available. Healthcare facilities are encouraged to use TAP to accelerate their prevention efforts. The CDC has developed several accessible spreadsheets for feedback reports, and there is also a Division of Healthcare Quality Promotion (DHQP) available to provide technical assistance for CAUTI prevention to facilities (Centers for Disease Control and Prevention [CDC], 2021).

Looking through the type of information submitted to the Journal of Infection Control & Hospital Epidemiology, several articles reflect upon the prevention of hospital-acquired infections (HAI). Beyond Bundles in Prevention of CAUTI and UTIs (see Appendix A) gives details of a CAUTI prevention project in a Texas healthcare facility. The project used “bladder bundles” which were closely monitored by a multidisciplinary quality improvement team. The project included safety rounds, staff education, quality audits, and consistent feedback. The project was widely adopted by the hospital system. There was a strong focus on looking for alternatives to indwelling foleys. Catheter days decreased from 162 to 49 by the completion of the project. The authors noted that the success of the project was due largely to the interprofessional team approach (Ford et al., 2020).

There was significant evidence supporting strict compliance to infection control policies. It is imperative that staff be properly trained on the avoidance, and proper use, when deemed necessary, of indwelling urinary catheters (Yeung, 2018). Medicare’s Partnership for Patients Initiative requested help from the American Nurses Association to help identify gaps in quality improvement efforts. Nurses from the WOCN (Wound, Ostomy, Continence Nursing) Society collaborated with other patient safety authorities to offer suggestions to complement or improve current evidence-based strategies. Together, they were able to develop a CAUTI prevention tool kit, which focused on the insertion and removal of indwelling foleys. It was piloted across 16 hospitals among medical, surgical, and oncology units (Lawrence et al., 2019). After reviewing the literature, the data was organized in a data matrix, which included the year of publication, study design, appraisal, and primary conclusions. Tools from the Joanna Briggs Institute (JBI) were used to evaluate levels of evidence.

## Chapter 4: Interventions

Project took place over an 8-week period from September to November 2021. To acquire and maintain stakeholder buy-in, the researcher requested the input of team members who have the most anticipated impact on the quality improvement initiative. It is important to identify current or past failed practices and contributing factors, discuss the current literature with the best evidence-based practices, and ultimately agree on a standardized intervention (Carr et al., 2019). The CAUTI GPS Assessment tool (see Appendix A) was used to survey and gauge the baseline knowledge of nursing staff, management, and providers. This tool was used as the formal needs assessment for the QI project. Specifically, the GPS tool was completed by one nurse manager, the assistant chief nursing officer, 3 medical-surgical nurses, and 11 critical care nurses.

Gaps in knowledge were analyzed, and then several weeks of education were streamlined to focus on CAUTI prevention processes. The researcher used section 1 of the Targeted Assessment for Prevention (TAP) tool (see Appendix B) which explored general infrastructure and processes of the facility’s CAUTI prevention efforts, section 2, which explored the appropriate indications for indwelling foleys, section 5, detailing timely removal of urinary catheters, and section 6 concerning appropriate urine culture practices. Healthcare workers who had a direct association with catheter use were surveyed.

Another crucial component of this QI project was to clarify facility criteria for situations when indwelling catheters are definitely indicated. Round table sessions with staff members provided the researcher an opportunity to assess whether nurses understood that there are unique situations in which indwelling foleys should be used. Meetings with key members of staff were held to explore ways to provide real-time data, visible to focus group members, staff, and providers related to current catheter usage, UTIs, and removals.

### Project Methodology

For this CAUTI (catheter-associated urinary tract infection) prevention project, the researcher has identified key personnel within the hospital system that can directly impact CAUTI rates. Charge nurses, nursing management, infection control managers, providers (i.e. physicians, nurse practitioners, physician assistants, surgeons), case managers, and information technology specialists are among the top stakeholders. The overarching aim is risk reduction, which benefits parties interested in patient safety, cost containment, and ultimately patient satisfaction.

The subjects in this quality improvement project included adult patients hospitalized in the medical surgical intensive care unit (ICU). This will include all patients (male and female) over the age of 18 on the unit. The participants will be nursing staff and nursing managers on the medical surgical units, specifically those in positions with unique or direct involvement with insertion, removal, or monitoring of foley catheters. Exclusion criteria (for the purpose of reviewing CAUTI cases) will include patients younger than 18.

### Evidence-Based Practice (EBP) Model

To promote sustainability of the QI project, the researcher will use Advancing Research & Clinical practice through close Collaboration (ARCC) Model as the evidence-based practice model. This model is initiated by evaluating the organizational culture and gauging the readiness for EBP. After identifying potential roadblocks, the researcher will collaborate with facilitators or change agents to obtain feedback, and ultimately buy-in. A plan will be formed to mitigate any barriers. Implementation of the ARCC Model leads to improved clinician’s knowledge and trust in their ability to promote real change, with improved outcomes for their patient populations (Tucker et al., 2021). If evidence-based practices are used consistently and reliably, up to 69% of CAUTI could be avoided (Fletcher et al., 2016).

### Nursing Theory

Kotter’s Change Model involves fostering cultural change through staff buy-in and persistent motivation and promotion of the QI initiative. A structured multidisciplinary team is well-prepared to effectively promote change which enhances the experiences of patients and improves outcomes. The use of Kotter’s model promotes leadership among nursing staff and prepares them to anticipate challenges and seek dynamic solutions. Kotter’s model contends that to achieve cultural change, the vision must be clear and shared (Mørk et al., 2018).

## Chapter 5: Results

### Data Analysis

The backbone of a quality improvement project is the measurement of outcomes. There must be a concerted effort to identify the benefit of change. Reasonable estimates of anticipated results should be clarified early in the project. To determine the success of the quality improvement project, several benchmarks will need to be identified along the way, with ultimate achievement of desired outcomes. A thorough review of the literature centered around the problem statement can help researchers find predictable outcomes. How the project impacts stakeholders is determined by proper outcomes measurement (Zaccagnini & White, 2017).

Survey results revealed that 87.5% of respondents felt that the facility’s leadership actively promoted CAUTI prevention. Only 43.7% believed that there was unit-level leadership involved in CAUTI prevention. 81% of respondents replied that the facility does have a team in place for CAUTI prevention, with 2 participants commenting “quality department.” Only 37.5% of respondents felt there was a specific nurse champion for CAUTI prevention activities. Every participant responded “no” when asked if there was a physician champion CAUTI prevention ins the facility. There were 4 comments left which indicated that the hospital has an infectious disease doctor on staff. 81.2% of participants responded yes when asked if senior leadership is supportive of CAUTI prevention activities. When asked about nurses assessing for continued need of indwelling foleys daily, 62.5% of participants responded yes. 87.5% of respondents answered yes when asked if they currently collect CAUTI-related data in the units. Over half of respondents felt that nurse resistance was a barrier to removing indwelling foleys. Only 37.5% of participants felt bedside nurses took initiative to remove indwelling foleys when no longer indicated. Over 81% of respondents reported receiving consistent feedback on CAUTI-related data.

It is imperative that strategies be utilized to protect the confidentiality of participants as well as the integrity of data when conducting formal research (Turcotte-Tremblay & Mc Sween-Cadieux, 2018). The researcher for this study has protected participants by utilizing a survey engine that does not reveal names of individual respondents. The data was secured on a passcode protected personal computer in a locked office. Data was entered by one researcher. To minimize error, the researcher double-checked all data entered prior to running statistical analyses.

## Findings

There was some nurse resistance to questionnaires, but most nurse leaders were receptive to questions and QI project in general. There were several reported gaps in staff knowledge related to roles of leadership personnel related to CAUTI prevention, based on survey feedback. The researcher then used the TAPS assessment recommendations to investigate further, to specifically place a focus on the immediate education needs. These sessions revealed that there was also insufficient knowledge related to when indwelling foleys were actually indicated. Most nurses felt ordering providers listed an indication or reason for ordering indwelling foleys. Bladder scans reportedly, are not consistently performed prior to insertion of urinary catheters. No frontline staff reported knowing whether there were standard protocols for acting on bladder scan results.

After modeling as champion for CAUTI prevention and obtaining buy-in throughout unit (and facility) for weeks, staff were more receptive. Several brief educational sessions were held during safety huddle and short round table type meetings. During these meetings, researcher was able to clarify current processes for prevention, number of indwelling foleys, number of non-invasive urinary drainage devices, indications for indwelling foleys, CAUTI occurrences, appropriate removals, advocating for alternatives to indwelling foleys (when appropriate), and discussing signs and symptoms of UTIs to monitor.

Identical survey questions were reissued at the end of the 8-week period. The independent t test was used to show differences between the pre and post phases with respect to a score. That score was derived by determining whether the participant answered a set of questions accurately. The scores represent the percentage answered correct. The independent t test is used when (1) you have two sub-populations with no common participants in both groups (2) an interval dependent variable, (3) the dependent variable is normally distributed. Interval variables are measurements based on a scale from 0 to infinity.

There is a statistically significant difference in scores between pre and post phase. There is a 22.5(5.8) mean difference between the two groups (t=-3.861, df=30, p<0.01)Results of the survey indicated that there was marked increase in staff awareness. Usage of non-invasive urinary drainage devices, such as Purewick Catheter systems, increased. There were no identified CAUTI during this period.

## Chapter 6: Conclusions

There must be an impetus to inspire compliance. If healthcare workers adhere to prevention guidelines, CAUTI are preventable. When leadership team members within hospital systems are enthusiastic about CAUTI prevention, the organization as a whole will have increased motivation (Chenoweth et al., 2014). After attending safety huddles and acting as a champion for CAUTI prevention, awareness of efforts, protocols, and indications improved.

Utilizing the CAUTI GPS Screening tool reduced the use of indwelling foleys in the in the inpatient setting. The use of alternative drainage devices, such as Purewick and condom catheters increased. Responses on repeat surveys from staff indicated that there was increased awareness of CAUTI prevention processes as well as increased staff accountability. There were no reported CAUTI during the QI period; however, the average number of catheter days did decrease.

### Limitations

There were several key limitations of this project. The main limitation was a low number of participants. The most profound roadblock was lack of participation or enthusiasm due to staffing shortage or nurse fatigue. The facility is burdened by nursing shortage during the COVID pandemic. Many team members were unmotivated to participate due to their workload. Many of the questionnaires were completed by travel nurses. While there were some full-time staff who completed the surveys prior to intervention, there was no ability for participant matching. Also, preliminary responses were questionable because several were completed by travel/agency nurses, who may have lacked baseline knowledge of the facility’s internal processes for infection control.

### Implications for Practice

It is imperative that there is a sound body of knowledge geared towards prevention, identification, and treatment of CAUTI. The benefits of prevention far outweigh the costs of implementing programs. Quality improvement processes should focus on reducing the use of indwelling catheters, when possible. Key stakeholders who stand to benefit from reduction in CAUTI are post surgical, elderly incapacitated patients, immunocompromised adults, clinicians, hospital administrators, and insurance companies.

Prevention of catheter-associated UTIs (CAUTI) is very important to scholarly nursing practice because it requires reinforcing and enhancing the knowledge of nurses at all level in the healthcare arena. Enhancing the overall patient experience and decreasing per capita costs per patient is the responsibility of all healthcare providers. The advanced practice nurse will play a lead role in the research, education, data collection, dissemination, and evaluation of the quality improvement project.

Advance practiced registered nurses (APRNs) pursing doctoral level education, must assure there is a strong focus on patient safety and standards of quality throughout their curriculum. This includes (but is not limited to) evaluation of care to identify gaps in quality, implementing indicated quality improvement projects, and faculty enrichment. When paired with quality tools and appropriate mentorship, doctoral level nurses can lead quality improvement teams throughout healthcare systems, and ultimately improve care globally (Kesten & Echevarria, 2021). The corner stone of competency for healthcare professionals is incorporating evidence-based practice into decisions. This will lead to increased patient satisfaction and improved health outcomes (Saunders et al., 2019).

## Data Availability

All data produced in the present work are contained in the manuscript.

## Appendix A

### CAUTI GPS Assessment Tool

#### CAUTI Guide to Patient Safety Tool

The catheter-associated urinary tract infection (CAUTI) Guide to Patient Safety (GPS) is a brief troubleshooting tool to aid infection prevention teams in reducing CAUTI in their hospital or unit.

Over the past decade, our multi-disciplinary research team has sought to better understand why some hospitals are more successful than others in preventing device-associated infection. This work includes conducting qualitative assessments of a total of 43 hospitals across the United States. In total, we have conducted 400 interviews of personnel at various levels within the organizations, from chief executive officers to front-line nurses and physicians.

From these interviews, we found that a handful of critical issues seemed to arise irrespective of a hospital’s location or size. These ranged from technical issues (e.g., collecting catheter use data) to common barriers to effective CAUTI prevention (e.g., lack of a physician champion). Because in person visits are both time-consuming and resource-intensive, we developed this self-administered list of questions that could be completed by key informants to help guide their hospital’s approach to CAUTI prevention.

#### Instructions for Use

To accurately assess the team’s CAUTI prevention efforts, it is recommended that:

1. The team working on CAUTI prevention at the hospital or unit-level completes the CAUTI GPS assessment. This can be done independently or as a group.
2. The responses are reviewed as a team as a means to uncover strengths and barriers to reducing CAUTI.
3. For questions 1 through 9 that were answered with a “No” and any part of question 10 that was answered “Yes,” the team should click on the link below the question or reference the indicated section to review approaches, advice, tools and resources to better implement the indicated CAUTI prevention strategy.

This tool was developed by faculty and staff from the Department of Veterans Affairs and the University of Michigan using funding support from the Department of Veterans Affairs, the University of Michigan, and the National Institutes of Health (NIH). This tool was validated and disseminated using funding support from the Agency for Healthcare Research and Quality (AHRQ), the Department of Veterans Affairs, and the University of Michigan.

#### CAUTI Guide to Patient Safety

##### Hospital_________________ Unit_________________

1. **Do you have a well-functioning team (or work group) focusing on CAUTI prevention?** ☐ Yes ☐ No *If you answered ‘No’ to the question above, review guidance and resources on having a <u>wellfunctioning team</u>*.
2. **Do you have a team leader with dedicated time to coordinate your CAUTI prevention activities?** ☐ Yes ☐ No *If you answered ‘No’ to the question above, review guidance and resources on having a <u>CAUTI team leader</u>*.
3. **Do you have an effective nurse champion for your CAUTI prevention activities**? ☐ Yes ☐ No *If you answered ‘No’ to the question above, review guidance and resources on <u>nurse champions</u>*.
4. **Do bedside nurses assess, at least daily, whether their catheterized patients still need a urinary catheter?** ☐ Yes ☐ No *If you answered ‘No’ to the question above, review guidance and resources on having a <u>daily assessment</u>*.
5. **Do bedside nurses take initiative to ensure the indwelling urinary catheter is removed when the catheter is no longer needed (e.g., by contacting the physician or removing the catheter per protocol)?** ☐ Yes ☐ No *If you answered ‘No’ to the question above, review guidance and resources on <u>removing unnecessary urinary catheter</u>*.
6. **Do you have an effective physician champion for your CAUTI prevention activities?** ☐ Yes ☐ No *If you answered ‘No’ to the question above, review guidance and resources on <u>physician champions</u>*.
7. **Is senior leadership supportive of CAUTI prevention activities?** ☐ Yes ☐ No *If you answered ‘No’ to the question above, review guidance and resources on <u>engaging senior leaders</u>*. **CAUTI Guide to Patient Safety** **Hospital**____________ **Unit**____________
8. **Do you currently collect CAUTI-related data (e.g., urinary catheter prevalence, urinary catheter appropriateness, and infection rates) in the unit(s) in which you are intervening?** ☐ Yes ☐ No *If you answered ‘No’ to the question above, review guidance and resources on <u>physician champions</u>*.
9. **Do you routinely feedback CAUTI-related data to frontline staff (e.g., urinary catheter prevalence, urinary catheter appropriateness, and infection rates)?** ☐ Yes ☐ No *If you answered ‘No’ to the question above, review guidance and resources on <u>engaging senior leaders</u>*.
10. **A. Have you experienced substantial nursing resistance?** ☐ Yes ☐ No *If you answered ‘Yes’ to the question above, review guidance and resources on <u>nursing resistance</u>*.
11. **B. Have you experienced substantial physician resistance?** ☐ Yes ☐ No *If you answered ‘Yes’ to the question above, review guidance and resources on <u>physician resistance</u>*.
12. **C. Have you experienced patient and family requests for an indwelling urinary catheter?** ☐ Yes ☐ No *If you answered ‘Yes’ to the question above, review guidance and resources on <u>engaging patients and families</u>*.
13. **D. Have you experienced indwelling urinary catheters commonly being inserted in the emergency department without an appropriate indication?** ☐ Yes ☐ No *If you answered ‘Yes’’ to the question above, review guidance and resources on <u>appropriate indications for urinary catheters</u>*.

### Question 1: Do you have a well-functioning team (or work group) focusing on CAUTI prevention?

**Because your catheter-associated urinary tract infection (CAUTI) prevention team is responsible for defining, designing, leading, and sustaining the initiative, it is crucial that it functions well**.

“For the change effort to be successful a powerful group must lead the change; and members of that group must work together as a team. Key characteristics that must be represented on the team include power, leadership skills, credibility, communications ability, expertise, authority, analytical skills, and a sense of urgency.” (From [TeamSTEPPS](https://www.ahrq.gov/teamstepps/index.html))

1. <u>Team Membership</u>
  - The composition of the team is important for the success of the initiative. We suggest that the team –at a minimum – include:
    - Team leader/project manager: When selecting a team leader, consider whether s/he has successfully led another quality improvement project. Leadership and management skills, and previous success are more important than the job title or content expertise.
  - Nurse champion: When selecting a nurse champion, consider someone who is well respected and in a position to obtain support from the other nurses given that avoiding catheter use may be perceived as additional nursing work (monitoring indwelling urinary catheter placement, increased toileting time, and possible data collection). We believe that having an effective nurse champion is critically important to the success of your initiative! For more information on overcoming a lack of/or challenges with a nurse champion, [click here](https://www.bladdersafe.org/resources.html). Select Engaging Providers tab and then Nurse Engagement. o Physician champion: When selecting a physician champion try to involve a physician who is highly regarded. If finding someone who is able to be actively engaged in the process is not possible, then consider selecting a respected physician who is willing to lend their name to this initiative. For overcoming a lack of/or challenges with a physician champion, [click here](https://www.bladdersafe.org/resources.html). Select Engaging Providers tab and then Physician Engagement.
  - Data person: Because the success of the intervention will be determined by the data, this person is a key component of any team. They are responsible for collating information-specifically, the presence of a Foley, the explanation for its original insertion or continued use, and any indication of a healthcare-associated urinary tract infection—and feed it back to the floor unit involved and to the hospital office responsible for sending the results to the CDC. This is often an infection preventionist, quality manager, or patient safety officer and it is common that s/he is already collecting and reporting the data to the internal leadership and for public reporting. For further information, see <u>Question 8</u>.
  - Other important team members can include a member of the senior leadership, a nurse educator, an infection preventionist, and a quality improvement officer.
  - Ideally the team will be composed of members with different backgrounds and various levels of experience.
  - Despite the possibility that the initiative may take place over many units, we suggest that there only be one CAUTI Prevention Team.
2. <u>What the Team Does</u> o The team must **take ownership** of the initiative. o Team members must **meet on a regular basis** (we suggest biweekly to begin).
  - They must **develop and implement an initiative**, which will involve educating healthcare providers of the existing evidence and severity of catheter complications.
  - They must **collect data** and feed it back to the unit.
3. <u>Information and Exercises for Team Evaluation and Improvement</u> o For a video overview of the assembly of a CAUTI prevention team, [click here](https://www.ahrq.gov/professionals/education/curriculum-tools/cusptoolkit/videos/02a_assemble_team/index.html). o For an example of a team assessment tool, please [click here](https://www.ahrq.gov/teamstepps/instructor/reference/tmassess.html).
  - For more information on team roles and responsibilities, [click here](http://www.catheterout.org/uploads/5/6/5/0/56503399/team_table.pdf).
4. <u>Further Reading Suggestions</u> o Damschroder LJ, Banaszak-Holl J, Kowalski CP, Forman J, Saint S, Krein SL. The role of the champion in infection prevention: results from a multisite qualitative study. Qual Saf Health Care.2009;18(6):434–40.
  - Fakih MG, Krein SL, Edson B, Watson SR, Battles JB, Saint S. Engaging healthcare workers to prevent catheter-associated urinary tract infection and avert patient harm. Am J Infect Control. 2014;42(10Suppl):S223-9.
  - Jain M, Miller L, Belt D, King D, Berwick DM. Decline in ICU adverse events, nosocomial infections and cost through a quality improvement initiative focusing on teamwork and culture change. Qual Saf Health Care.2006;15(4):235-9.

### Question 2: Do you have a team leader with dedicated time to coordinate your CAUTI prevention activities?

**Because the project manager (also referred to as the team leader) is responsible for coordinating and facilitating meetings, team communication, and overseeing that members understand their roles and follow through on their responsibilities, it is imperative that s/he has dedicated time to commit to the project**.

1. <u>If nobody has been identified for this role</u> o Ask senior leadership for advice about who they recommend and who they can provide with some protected time to do this work.
  - Find someone who has been successful in coordinating a quality improvement initiative.
  - Experience and knowledge of the topic is secondary in importance to leadership skills, enthusiasm, persistence, and credibility. The leader will be expected to reach out to the content experts for guidance related to the technical aspects of the work.

1. <u>If the selected project manager is not as effective as necessary</u> o Check to see if s/he has been given dedicated time to work on this particular project. If not, engage leadership to help with this.
  - S/he may be lacking some of the necessary skills. We have found that coaching him/her on what they can improve upon can be very helpful.
  - S/he may not be a good fit for the initiative, and it may be time to consider replacing him/her with someone else.
2. <u>For a better understanding of what makes a project manager successful</u> o [Top 10 Qualities of a Project Manager](https://www.projectsmart.co.uk/top-10-qualities-project-manager.php) o [Top 10 Characteristics of GREAT Project Managers](https://www.behance.net/blog/top-10-characteristics-of-great-project-managers)
3. <u>Further Reading Suggestions</u> o Cannon-Bowers, J. A., S. I. Tannenbaum, E. Salas, and C. E. Volpe. “Defining competencies and establishing team training requirements”. Team effectiveness and decision-making in organizations. Ed. R.A. Guzzo, E. Salas, and Associates: San Francisco: Jossey-Bass, (1995)333.
  - Salas E, Burke CS, Stagl KC. “Developing teams and team leaders: strategies and principles.” Leader Development for Transforming Organizations. Ed. R. G.Demaree, S. J. Zaccaro, and S. M. Halpin: Mahwah, NJ: Lawrence Erlbaum Associates, Inc, (2004).

### Question 3: Do you have an effective nurse champion for your CAUTI prevention activities?

**Because a CAUTI initiative is especially dependent on nurses, an effective nurse champion is key. By effective we mean: well respected and trusted by peers, supportive of safety, and an agent of change**.

1. <u>If a nurse champion is not yet identified</u> o The most successful nurse champions are those that know their way around the hospital hierarchy but are independent-minded in terms of finding solutions. S/he must be on good terms with her colleagues.
  - Think twice about having a nurse executive or the director of nursing take on this role as there is danger that the bedside nurses may view the initiative as another occasion for obeying the boss.
  - Some qualities that make a successful nurse champion include: being personable, enthusiastic, empathetic, and having great communication skills.
  - There is no “one-size-fits-all” strategy. You must identify the type of individual that will work best in your organization. o Consider a nurse manager, nurse educator, and even a respected licensed practical nurse who others go to for advice.
  - Consider having co-champions if necessary.
  - Far more than the physician champion, the nurse champion is the face of the project to the people most instrumental in the project’s success, the bedside nurses.
2. <u>If the nurse champion on your team is not as effective or engaged as needed</u> o As with other members of the team, we have found that often the nurse champion has not been given dedicated time to work on this particular project. Supporting the nurse champion (e.g., reducing some of the clinical commitments in order to address the quality improvement project) during the initial stages may help with implementation efforts.
  - If the nurse champion has the dedicated time but is lacking some of the necessary skills, we have found that coaching him/her can be very helpful. The ideal coach may be the senior leader who is tracking the outcome of this project.
  - It is important to choose the champion based on his/her commitment to the issue and interest in safety. If it is clear that the nurse champion is not a good fit for the initiative (s/he may have been appointed rather than recruited), it may be time to consider replacing the nurse champion.
  - It is also important to recognize the nurse champion for his/her efforts via such mechanisms as certificates of recognition, annual evaluation appraisals, mention in newsletters, and acknowledgement from the Chief Nursing Officer.
  - We have also found that identifying and enlisting others who are either already engaged in this initiative or eager to improve patient safety can help support the efforts of the nurse champion.
3. <u>For more information on nurse engagement</u>, [click here](http://www.catheterout.org/resources.html). *Select Engaging Providers tab and then Nurse Engagement*
4. <u>Further Reading Suggestions</u> o Draper, DA, Felland, LE, Liebhaber, A, Melichar, L. The role of nurses in hospital quality improvement. Research Brief No. 3, March 2008; Center for Studying Health System Change. http://www.hschange.com/CONTENT/972/972.pdf o Gokula RM, Smith MA, Hickner J. Emergency room staff education and use of a urinary catheter indication sheet improves appropriate use of Foley catheters. Am J Infect Control.2007;35(9):589-93.

### Question 4: Do bedside nurses assess, at least daily, whether their catheterized patients still need a urinary catheter?

**Because the necessity of an indwelling urinary catheter may change while a patient is in the hospital, it is imperative to continually assess its appropriateness. Daily assessment of catheter necessity is perhaps the single most important method of decreasing catheter use and subsequent infection**.

1. <u>Remind/educate your staff</u> o Urinary catheters are often placed unnecessarily, remain in place without physician awareness, and are not removed promptly when no longer needed.
  - Prolonged catheterization is the strongest risk factor for catheter-associated urinary tract infection (CAUTI).
  - Promptly removing unnecessary catheters is an important step in reducing a patient’s risk of CAUTI.
2. <u>Indwelling urinary catheters should be addressed daily</u> o If nurses are concerned that they will have to spend more time cleaning up patients if the indwelling urinary catheter is removed, try:
  - Timed voiding or hourly intentional rounding
  - Exploring incontinence products, urinals, condom catheters, and intermittent straight catheters
  ∘ If there is a general feeling of being overworked (“just trying to get through my shift”), try:
  - “Catheter patrol” - One or two daytime charge nurses who monitor which patients have indwelling urinary catheters, assist with toileting, and assess the indications for urinary catheters. If the indwelling urinary catheter is no longer clinically indicated, the “catheter patrol” can talk with the bedside nurse or ask the physician directly to discontinue.
  - Daily assessment tool - Tailored to the care setting, bedside nurses (or the “catheter patrol”) can assess the indications for the continued use of indwelling urinary catheters and if no longer clinically indicated, nurses can discuss its removal with the physician. Click here for an example.
3. If there is no mechanism to trigger prompt removal, consider:
  - Stop orders that prompt catheter removal by default after a certain time period or a set of clinical conditions has occurred (such as 24 or 48 hours postoperative) unless the catheter remains clinically appropriate. **Or**
  - A nurse-initiated removal protocol—whereby a nurse can initiate the removal of the indwelling urinary catheter by contacting the physician if after assessment an indication for continued use has not been identified.
4. <u>For more information</u> o For the current appropriateness guidelines, see the [Ann Arbor Criteria](http://www.catheterout.org/ann-arbor-criteria.html).
  - For more information and examples of nurse driven protocols to evaluate and discontinue unnecessary urinary catheters, [click here](https://bladdersafe.org/uploads/5/6/5/0/56503399/example_of_a_nurse_driven_protocol_mgf_6-2-12.pdf).
  - For more information on daily evaluation of urinary catheter appropriateness, see the information under <u>Question 8</u>.
5. <u>Further Reading Suggestions</u> o Elpern EH, Killeen K, Ketchem A, Wiley A, Patel G, Lateef O. Reducing use of indwelling urinary catheters and associated urinary tract infections. Am J CritCare.2009;18(6):53541; quiz42.
  - Fakih MG, Watson SR, Greene MT, Kennedy EH, Olmsted RN, Krein SL, Saint S. Reducing inappropriate urinary catheter use: a statewide effort. Arch Intern Med. 2012;172(3):255-60.
  - Fakih MG, Pena ME, Shemes S, et al. Effect of establishing guidelines on appropriate urinary catheter placement. Acad Emerg Med.2010;17:337-40.
  - Fakih MG, Dueweke C, Meisner S, et al. Effect of nurse-led multidisciplinary rounds on reducing unnecessary use of urinary catheterization in hospitalized patients. Infect Control Hosp Epidemiol.2008;29:815–9.
  - Fuchs MA, Sexton DJ, Thornlow DK, Champagne MT. Evaluation of an evidence-based, nurse-driven checklist to prevent hospital-acquired catheter-associated urinary tract infections in intensive care units. J Nurs Care Qual.2011;26(2):101-9.
  - Gokula RR, Hickner JA, Smith MA. Inappropriate use of urinary catheters inelderly patients at a midwestern community teaching hospital. Am J Infect Control. 2004;32:196-9.
  - Meddings J, Rogers MA, Krein SL, Fakih MG, Olmsted RN, Saint S. Reducing unnecessary urinary catheter use and other strategies to prevent catheter-associated urinary tract infection: an integrative review. BMJ Qual Saf.2014;23(4):277-89.
  - Meddings J, Rogers MA, Macy M, Saint S. Systematic review and meta-analysis:
  - reminder systems to reduce catheter-associated urinary tract infections and urinary catheter use in hospitalized patients. Clin Infect Dis.2010;51:550-60.
  - Miller BL, Krein SL, Fowler KE, et al. A multimodal intervention to reduce urinary catheter use and associated infection at a Veterans Affairs Medical Center. Infect Control Hosp Epidemiol. 2013;34(6),631–633.
  - Saint S, Wiese J, Amory JK, et al. Are physicians aware of which of their patients have indwelling urinary catheters? Am J Med.2000;109:476-80.

### Question 5: Do bedside nurses take initiative to ensure the indwelling urinary catheter is removed when the catheter is no longer needed (e.g., by contacting the physician or removing the catheter per protocol)?

**Because timely removal of the indwelling urinary catheter is crucial for reducing catheter-associated urinary tract infection (CAUTI), nurses should be empowered and supported to take the initiative to remove the catheter when it is no longer appropriate (e.g., by contacting the physician or removing the catheter per approved protocol)**.

1. <u>Policy to trigger prompt removal is key</u> o Stop orders which prompt the clinician to remove the catheter by default after a certain time period or a set of clinical conditions has occurred (such as 24 or 48 hours post-operative) unless the catheter remains clinically appropriate.
  - Stop orders “expire” in the same fashion as restraint orders or antibiotic orders, unless action is taken by physicians.
  - Urinary catheter reminders simply alert doctors and bedside nurses to the fact that a Foley is being used by a patient and provide a list of the appropriate reasons to continue or discontinue the indwelling catheter.
  - Reminders are generally dispatched as a hospital unit eases into an infection prevention initiative.
  - The reminder is included in the patient’s chart or is part of the patient’s electronic record.
  - The use of <u>daily appropriateness tracking</u> can be helpful for decreasing unnecessary indwelling urinary catheters. Bedside nurses make a daily entry indicating whether any given Foley meets one or more of the appropriate indications for catheter use. If an inplace catheter fails that test, the nurse is to alert the appropriate physician caring for the patient and recommend the catheter’s removal. Additional information can be found under <u>Question 8</u>.
  - Some hospitals have had great success with a [nurse-initiated removal protocol](https://www.bladdersafe.org/detailed-protocols.html) whereby a bedside nurse can initiate the removal of the indwelling urinary catheter without an attending physician order; however, this usually needs to be approved by a Medical Executive Committee first, and should be presented by a physician.
2. <u>Resistance to early Foley removal, a common barrier</u> o Educate staff members
  - Urinary catheters are often placed unnecessarily, remain in place without physician awareness, and are not removed promptly when no longer needed. Prolonged catheterization is the strongest risk factor for CAUTI.
  - For more information on infectious complications, [click here](https://www.bladdersafe.org/resources.html), then click Infectious Complications.
  - For examples of power point presentations, [click here](https://www.bladdersafe.org/resources.html). *Select the Educational Tools tab and then click Presentations*.
  - Enlist champions and supporters
  - When physicians, especially urologists and surgeons are resistant, have the physician champion present information about the indications and nonindications for the indwelling urinary catheter at a medical staff meeting. ? Engage a surgeon and/or urologist as a physician champion and work with them to establish conditions under which the catheter can be removed. ? Work with physician assistants or nurse practitioners to discontinue Foleys within 1 or 2 days after surgery
  - Identify and promote other benefits to catheter removal
  - Earlier mobility
  - Decreased non-infectious complications (urine leakage, gross hematuria and urethral strictures)
  - Earlier discharge potential
3. <u>Challenges and pearls to keep in mind when implementing catheter removal strategies</u> o Capitalize on “nurse-to-nurse” communication at times of care transition (between shift and between units) as opportunities to reassess catheter need. Having a nurse champion on every shift may facilitate reassessment, especially if shift schedules make it difficult to share information.
  - Simple reminders are often ignored. It can be challenging to sustain the impact of reminders.
  - Reminder system chosen should be tailored to the care setting (stickers, electronic, etc.). Both low-tech and high-tech strategies have been effective.
  - If using electronic reminders/stop orders, make sure the reminder/stop order is directed at the primary team and not the consultants. o Using electronic catheter orders can increase catheter use inadvertently by making indwelling catheters easier to order than alternatives.
  - Physicians and/or nurses should document the rationale for leaving the catheter in if appropriate indications are not met. Documentation makes the rationale explicit and communicates it to the rest of the healthcare team.
  - Nurses may not be comfortable initially with the responsibility of removing urinary catheters without a physician order. Supportive nursing and physician leadership can help overcome nurses’ reluctance to act.
  - Incontinence is a very tempting reason for placing a urinary catheter. Encourage bedside staff to avoid placing catheters for incontinence by providing other readily available strategies to manage incontinent patients, including bedside commodes, incontinence garments, condom catheters for male patients, and “people power” to provide prompted toileting and bed linen changes.
4. <u>Further Reading Suggestions</u> o Fakih MG, Rey JE, Pena ME, Szpunar S, Saravolatz LD. Sustained reductions in urinary catheter use over 5 years: bedside nurses view themselves responsible for evaluation of catheter necessity. Am J Infect Control. 2013;41(3):236-9.
  - Fink R, Gilmartin H, Richard A, Capezuti E, Boltz M, Wald H. Indwelling urinary catheter management and catheter associated urinary tract infection practices in Nursing
  - Improving Care for Healthsystem Elders Hospitals. Am J Infect Control.2012; 40:715-20. o Meddings J, Rogers MA, Krein SL, Fakih MG, Olmsted RN, Saint S. Reducing unnecessary urinary catheter use and other strategies to prevent catheter-associated urinary tract infection: an integrative review. BMJ Qual Saf.2014;23(4):277-89.
  - Meddings J, Rogers MAM, Macy M, Saint S. Systematic review and meta-analysis:
  - reminder systems to reduce catheter-associated urinary tract infections and urinary catheter use in hospitalized patients. Clin Infect Dis.2010;51(5):550-60.
  - Oman KS, Makic MB, Fink R, SchraederN, Hulett T, Keech T, Wald H. Nurse-directed interventions to reduce catheter-associated urinary tract infections. Am J Infect Control.2012;40:548-53.
  - Patrizzi K, Fasnacht A, Manno M. A collaborative, nurse-driven initiative to reduce hospital-acquired urinary tract infections. J Emerg Nurs.2009;35:536-9.

### Question 6: Do you have an effective physician champion for your CAUTI prevention activities?

**Because catheter-associated urinary tract infection (CAUTI) prevention efforts require collaboration and support of both physicians and nurses, an effective physician champion can be important**.

1. <u>To identify a physician champion</u> o There is no “one-size-fits-all” strategy. You must identify the type of physician that will work best in your organization. Some suggestions include hospital epidemiologists, hospitalists, infectious diseases specialists, and urologists. Beware of choosing people on the basis of their job title. Unfortunately, titles don’t guarantee that a person will be appropriate for this task.
  - Our experience has been that the most successful physician champions are those that have pride in the hospital’s culture of excellence or concern over the lack of one. Ideally they are ideally a person who has the ear of the hospital administration and the respect of his or her peers, a doctors’ doctor, and someone who has the patience to hear out people who disagree with his or her point of view.
  - Because many physicians are not employees of the hospital and convincing a physician, employee or not, to take on any extra work is likely a tough assignment, we suggest the following:
    - While we do not believe that paying doctors to take part in a patient-centered intervention is necessary or preferred, we see no problem with
      - Temporarily relieving the physician of some of his/her responsibilities
      - Or as was done in one hospital, recognizing a member of the medical staff with a “physician champion” award, complete with a certificate signed by the hospital’s chief of staff and a gift certificate to a local restaurant.
    - Assure the physician champion that their role will not take too much of their time. They should not, for example, be expected to attend all meetings or be otherwise involved in matters unrelated to clinical concerns such as budget discussions or internal promotional plans or working out details of data collection, unless of course they want to be. Their chief responsibility will be to share the details of the intervention with colleagues and gain their cooperation.
2. <u>If the physician champion on your team is not as effective or engaged as needed</u> o In institutions where there are good nurse-physician working relationships, most physicians may be willing to go along with recommendations by nurses, especially if the new practice is viewed as a “nursing initiative.”
  - As with other members of the team, we have found that in many instances the physician champion has not been given dedicated time to work on this particular project.
  - Make sure that medical leadership supports the initiative.
  - A ‘strong’ physician champion may not be entirely necessary if both nurses and medical leadership supports the initiative and there is no active resistance from physicians. o Find a member of the ‘tribe’. “Surgeons are very tribal,” the chief of staff said, discussing the difficulty an infection prevention leader (an internist) might have trying to bring his message to a group of surgeons. “The first thing we’re going to do is we’re going to say, ‘Look, you’re not one of us.’ The way to get buy-in from surgeons is you got to have a surgeon on your team.”
3. <u>Further Reading Suggestions</u> o Damschroder LJ, Banaszak-Holl J, Kowalski CP, Forman J, Saint S, Krein SL. The role of the champion in infection prevention: results from a multisite qualitative study. Qual Saf Health Care.2009;18(6):434-40.
  - Fakih MG, Krein SL, Edson B, Watson SR, Battles JB, Saint S. Engaging healthcare workers to prevent catheter-associated urinary tract infection and avert patient harm. Am J Infect Control. 2014;42(10Suppl):S223-9.
  - Reinertsen JL, Gosfield AG, Rupp W, Whittington JW. Engaging physicians in a shared quality agenda. IHI Innovation Series white paper. (2007);Cambridge, MA: Institute for Healthcare Improvement. (Available on www.IHI.org) o Saint S, Kowalski CP, Banaszak-Holl J, Forman J, Damschroder L, Krein SL. The importance of leadership in preventing healthcare-associated infection: results of a multisite qualitative study. Infect Control Hosp Epidemiol.2010;31(9):901-7.

### Question 7: Is senior leadership supportive of CAUTI prevention activities?

**It is helpful if hospital administrators and clinical chiefs take on personal leadership roles in quality improvement initiatives. With some extra effort they can help build powerful support for the catheter-associated urinary tract infection(CAUTI)prevention project. Ideally, one member of the executive leadership team will be primarily responsible for overseeing the CAUTI initiative at your hospital. In our experience, this often is the chief nursing executive**.

1. <u>To engage leadership</u> o Prepare and present a business case to help convince leadership that the time and resources for implementing the new practice will be worth it.
  - The [CAUTI Cost Calculator](https://www.bladdersafe.org/cauti-cost-calculator.html) estimates your hospital’s costs due to CAUTI. It can be used to estimate both the current and projected costs after a hypothetical intervention to reduce catheter use. o Be sure leadership receives monthly CAUTI rates and catheter use data.
2. <u>Ways that leadership can show their support</u> o Mention in meetings and other staff encounters that these prevention activities are a reflection of the hospital’s mission
  - Stop by and listen in to a reporting session on the initiative, thus boosting the team’s sense of purpose;
  - Include updates on the project’s progress in their hospital-wide newsletter and online communications;
  - Make the degree of a person’s support of quality initiatives a regular element of employee performance reviews;
  - Top supervisors can provide backing when those leading an initiative run up against immovable road blocks.
3. <u>Further Reading Suggestions</u> o Kotter J. Leading change: why transformation efforts fail. Harv Bus Rev.1995;59-67. o Saint S, Kowalski CP, Banaszak-Holl J, Forman J, Damschroder L, Krein SL. The importance of leadership in preventing healthcare-associated infection: results of a multisite qualitative study. Infect Control Hosp Epidemiol.2010;31:901-7.

### Question 8: Do you currently collect CAUTI-related data (e.g., urinary catheter prevalence, urinary catheter appropriateness, and infection rates) in the unit(s) in which you are intervening?

**Collecting and comparing data both before and after an intervention provides an objective way to evaluate if your interventions are successful in reducing unnecessary catheter days and catheterassociated urinary tract infection (CAUTI). Ongoing assessments allow you to assess if the intervention is sustained**.

1. <u>The what and when of data collection</u> o <u>What to collect:</u>
  - The presence of a Foley
  - The explanation for its original insertion or continued use
  - Number of symptomatic CAUTI o <u>When to collect it:</u>
  - At baseline: daily for 2 weeks (phase1)
  - During implementation: daily for two weeks (phase two) ? After implementation: one day a week for 5 weeks (phase3) ? During sustainability: daily for one week each quarter (phase4)
2. <u>Calculations you should make from the data you collect:</u>
  - Process measure:
    - Catheter utilization rate: Total # catheter-days/Total # patient-days X100 o Outcome measure:
    - [NHSN measure](https://www.cdc.gov/nhsn/enrolled-facilities/index.html): # of symptomatic CAUTI/1,000 urinary catheter days as measured in NHSN.
    - Population-based measure: Total # of symptomatic CAUTI/10,000 patient days o Additional measures to consider
    - Unnecessary Urinary Catheter %: # of unnecessary catheter-days/Total # catheter-days X100
  - For more information on these calculations [click here](https://bladdersafe.org/uploads/5/6/5/0/56503399/data_collection___evaluation_detailed_2.pdf).
3. <u>It is important to apply a consistent approach to data collection at all stages of your prevention program so that you can compare across time periods and units.</u>
  - For an example of a data collection tool <u>click here</u>. Feel free to use or modify this in any way.
4. <u>Ensure that you have someone on the team who is responsible for collecting data</u> o This is typically an infection preventionist or a member of the quality improvement department.
  - Responsibilities of this team member include
  - Collecting and collating information –specifically, the presence of a Foley, the explanation for its original insertion or continued use, and any indication of a healthcare-associated urinary tract infection.
  - CAUTI Guide to Patient Safety (GPS) Q<u>8www.</u>c<u>atheterout.org2Last</u> updated 10/3/2014 Feeding it back to the floor unit involved and to the hospital office responsible for sending the results to the CDC.
5. <u>Further Reading Suggestions</u> o Choudhuri JA, Pergamit RF, Chan JD, et al. An electronic catheter-associated urinary tract infection surveillance tool. Infect Control Hosp Epidemiol.2011;32(8):757-62.
  - Fakih MG, Greene MT, Kennedy EH, et al. Introducing a population-based outcome measure to evaluate the effect of interventions to reduce catheter-associated urinary tract infection. Am J Infect Control.2012;40(4):359-64.
  - Trick WE, Samore M. Denominators for device infections: who and how tocount.Infect Control Hosp Epidemiol. 2011;32(7):641-3. o Wright MO, Kharasch M, Beaumont JL, Peterson LR, Robicsek A. Reporting catheterassociated urinary tract infections: denominator matters. Infect Control Hosp Epidemiol.2011;32(7):635-40.
6. For an example data collection process currently used by several hospitals [click here](https://bladdersafe.org/uploads/5/6/5/0/56503399/data_collection___evaluation_detailed_2.pdf).

### Question 9: Do you routinely feedback CAUTI-related data to frontline staff (e.g., urinary catheter prevalence, urinary catheter appropriateness, and infection rates)?

**Actively communicating with and providing timely and useful feedback to staff is an important part of quality improvement. Many hospitals have found that the transparency of sharing as much information as possible with the staff can help staff stay motivated and engaged in the quality improvement initiative**.

1. <u>Data that should be fed back to frontline staff</u> o What to collect: frontline staff o Data from the intervention o Any comparable data from nearby [hospitals](https://www.medicare.gov/hospitalcompare/search.html?AspxAutoDetectCookieSupport=1) o Any comparable national [data](https://www.cdc.gov/nhsn/)
2. <u>Mechanisms for feedback</u> o “Scorecard” that provides information on how performance is progressing toward goals
  - Can be provided at both the hospital and unit level
  - Should be visibly displayed throughout the hospital for all staff to see o Newsletters o Staff training o New employee orientation o e-mail Communications o Staff meetings
3. <u>The key to effective feedback is not just the amount of information provided, but also how meaningful that information is for staff.</u>
  o Do not limit feedback to numbers (e.g., CAUTI rate) provide details to make it more meaningful to the staff (e.g., we have gone X days since our last CAUTI).
4. <u>Rewarding the staff or a unit for positive changes can be motivating</u> o For example:
  - One site gave a little treat when a nurse initiated an early removal of a urinary catheter
  - Another site provided a pizza party to a unit that was able to get their high CAUTI rate down to zero
5. <u>Further Reading Suggestions</u> o Dubbert PM, Dolce J, Richter W, Miller M, Chapman SW. Increasing ICU staff handwashing: effects of education and group feedback. Infect Control HospEpidemiol. 1990;11(4):191-3.

### Question 10A: Have you experienced substantial nursing resistance?

**In a catheter-associated urinary tract infection (CAUTI) prevention program, the nursing staff, especially frontline staff, are central to the success of the initiative. Because they are the staff whose day-to-day activities are most affected by the changes, they may present the greatest resistance**.

1. <u>Reason for the resistance</u> o Because resistance can occur for a number of different reasons, as a first step we suggest interviewing front-line staff to learn why they are resistant to implementing a CAUTI prevention program and what, in the opinion of staff, is needed before acceptance of the program can occur.
2. <u>Strategies for enhancing nursing engagement and decreasing potential resistance</u> o Get a volunteer from the nursing staff to be a change champion for each shift— someone who other staff respect and who is committed to the process (examples include a front line nurse or a nurse educator).
  - Get buy-in before implementation. For example, ask, “Whom do we have to convince on this floor?” Have that person help to develop the plan and/or participate in the education for that unit. o Provide regular feedback on progress, as well as monthly reports on urinary catheter prevalence, and CAUTI rates. o Encourage nurses to be creative, developing visual cues to stimulate interest and keep the CAUTI initiative a top priority.
    - One site posted flyers/banners on the unit, such as “This is a catheter out zone.”
  - Make sure to listen and clearly understand nurses’ concerns and address them to the nurses’ satisfaction. This may require some education of the staff, creativity, or reallocation of resources.
  - Consider changes to (or redistribution of) workload.
    - For example, one site instituted a “small zone” so that nurses could be given a somewhat lighter workload when assigned to a patient who needed help with frequent toileting.
    - Another strategy is to prioritize nurse assistant/tech tasks to toileting patients.
  - Bring the education to the bedside. Do competencies on the unit, talking with nurses one-to-one during the point prevalence assessments.
3. <u>For more information</u> o On nurse engagement click [here](http://www.catheterout.org/resources.html). Select the Engaging Providers tab, and then Nurse Engagement. o On barriers and solutions click [here](http://www.catheterout.org/resources.html). Select Engaging Providers tab, then Barriers and Possible Solutions.
4. <u>Further Reading Suggestions</u> o Krein SL, Kowalski CP, Harrod M, Forman J, Saint S. Barriers to reducing urinary catheter use: a qualitative assessment of a statewide initiative. JAMA Intern Med. 2013;173(10):881-6. o CAUTI Guide to Patient Safety (GPS) Q<u>10Aw</u>ww<u>.c</u>a<u>theterout.org2Last</u> updated 10/3/2014oSaint S, Kowalski CP, Banaszak-Holl J, Forman J, Damschroder L, Krein SL. The importance of leadership in preventing healthcare -associated infection: results of a multisite qualitative study. Infect Control Hosp Epidemiol.2010;31:901-7. o Saint S, KowalskiCP, Banaszak-Holl J, Forman J, Damschroder L, Krein SL. How active resisters and organizational constipators affect health care-acquiredinfection prevention efforts. Jt Comm J Qual Patient Saf.2009;35:239-46.
  ∘ Saint S, Kowalski CP, Forman J, et al. A multicenter qualitative study on preventing hospital-acquired urinary tract infection in US hospitals. Infect Control Hosp Epidemiol. 2008;29:333-41.

### Questions 10B: Have you experienced substantial physician resistance?

**Because the day-to-day operation of a quality improvement project requires the ability of staff to adopt new goals and practices, it is important that the physicians either embrace, or at a minimum do not resist the implementation of catheter-associated urinary tract infection (CAUTI) prevention activities at your site/unit**.

1. <u>If there are some physicians who are resisting the initiative</u> o Educate them on the clinical and economic consequences of continuing the status quo.
  - Clinical consequences are both infectious and non-infectious. [Click](http://www.catheterout.org/resources.html) here for more information on clinical consequences. There are sections for both Infectious or Non-infectious Complications.
  - The [CAUTI Cost Calculator](https://www.bladdersafe.org/cauti-cost-calculator.html) estimates your hospital’s costs due to CAUTI. It can be used to estimate both current costs and projected costs after a hypothetical intervention to reduce catheter use.
2. Provide data to physicians about Foley use highlighting
  - how often physicians have a patient with an indwelling urinary catheter and forget about
  - it monthly Foley incidence
  - CAUTI rates o Engage medical leadership support by discussing the issue of CAUTI with the chief of staff (or chief medical officer) who in turn can, as needed, have a frank conversation with physician resistors. o Involve the physicians as much as possible in the planning, education, and implementation of the project.
3. Identify and discuss specific reasons why catheter use might be of interest for a given type of physician.
  - For example, a geriatrician might be inclined to support catheter removal given that urinary catheters increase immobility and is a deconditioning risk for their already frail patients.
4. If you are still struggling with CAUTI efforts related to physician engagement, it may be useful to determine the type of people-related issues you may be confronting: active resistance, organizational constipation, and time-serving.
  - For more information related to this [click here](http://www.catheterout.org/uploads/5/6/5/0/56503399/styles_of_personnel_barriers.pdf).
5. <u>For more specific suggestions for engaging physicians</u>, [click here](https://www.bladdersafe.org/resources.html). Select Engaging Providers tab and then Physician Engagement.
6. <u>For existing presentations, fliers, and pocket cards</u>, [click here](https://www.bladdersafe.org/resources.html). Select Educational Tools tab and then Presentations.
7. <u>Further Reading Suggestions</u> o Fakih MG, Rey JE, Pena ME, Szpunar S, Saravolatz LD. Sustained reductions in urinary catheter use over 5 years: bedside nurses view themselves responsible for evaluation of catheter necessity. Am J Infect Control. 2013;41(3):236-9.
  - Dyc NG, Pena ME, Shemes SP, Rey JE, Szpunar SM, Fakih MG. The effect of resident peer-to-peer education on compliance with urinary catheter placement indications in the emergency department. Postgrad Med J.2011;87(1034):814-8.
  - Kalra R, Kraemer RR. LESS IS MORE Urinary Catheterization—When Good Intentions Go Awry A Teachable Moment. JAMA Intern Med. Published Online: August 18, 2014.doi:10.1001/jamainternmed.2014.3806.
  - Kennedy EH, Greene MT, Saint S. Estimating hospital costs ofcatheter-associated urinary tract infection. J Hosp Med2013;9(9):519-522.oSaint S, Wiese J, Amory JK, et al. Are physicians aware of which of their patients have indwelling urinary catheters? Am J Med.2000;109:476-80.
  - Umscheid CA, Mitchell MD, Doshi JA,Agarwal R, Williams K, Brennan PJ. Estimating the proportion of healthcare-associated infections that are reasonably preventable and the related mortality and costs. Infect Control Hosp Epidemiol.2011;32(2):101-14.
8. For an example of one hospital’s success at overcoming this barrier, [click here](https://bladdersafe.org/uploads/5/6/5/0/56503399/example_of_overcoming_physician_lack_of_engagement.pdf).

### Question 10C: Have you experienced patient and family requests for an indwelling urinary catheter?

**Educating patients and their family members about the importance of urinary catheter risks can be an important way to reduce the unnecessary use of urinary catheters**.

1. <u>Patients and families may believe that the use of indwelling urinary catheters is in the patient’s best interest, however we have found that this is often based on incomplete, and sometimes incorrect, information. It is important to educate your patients and their family members about the risks of a Foley, the benefits of early removal, and alternative toileting options.</u>
  - Prepare and present a business case to help convince leadership that the time and resources for implementing the new practice will be worth it.
  - Written by members of our team, <u>“What Patients and Family Members Need to Know About the Risks Associated with Urinary Catheters”</u> is a brochure that offers information about urinary catheters, appropriate indications, and ways to discuss its use. This brochure can be tailored to your site.
  - Distributed by the Society for Healthcare Epidemiology of America, the one-page sheet, [“FAQs about Catheter-Associated Urinary Tract Infections”](https://bladdersafe.org/uploads/5/6/5/0/56503399/tab_p_part_iii_cauti_for_patients_shea.pdf), provides patients with an overview of urinary catheters, catheter-associated urinary tract infections, and how patients can safely care for their urinary catheter.
  - It may be helpful to practice various scenarios. See [“Script for patient or family requests for non-medically indicated indwelling urinary catheters”](https://bladdersafe.org/uploads/5/6/5/0/56503399/patient_and_family_script_final_1.pdf) for suggestions of statements that we have found useful
2. <u>For other resources, please visit:</u> o http://www.ahrq.gov/professionals/systems/hospital/engagingfamilies/index.html o http://www.rwjf.org/en/research-publications/find-rwjf-research/2013/02/patientengagement.html
3. <u>Further Reading Suggestions</u> o Krein SL, Kowalski CP, Harrod M, Forman J, Saint S. Barriers to reducing urinary catheter use: a qualitative assessment of a statewide initiative. JAMA Intern Med. 2013;173(10):881-6

### Question 10D: Have you experienced indwelling urinary catheters commonly being inserted in the emergency department without an appropriate indication?

**In the hectic and unpredictable environment of the emergency department (ED), physicians and nurses properly see themselves as serving on the front lines. Nurses and doctors are more concerned about whether their patients are still breathing than about whether they have a catheter. It takes a member of the catheter-associated urinary tract infection (CAUTI) prevention team to convince the ED that catheters count**.

1. <u>Indwelling urinary catheters are commonly placed automatically in the ED</u> o The CAUTI prevention team should include emergency department personnel (e.g., emergency medicine physician and nurse) when the initiative moves to the ED. o With the emergency medicine physician leading the way, the ED staff should be convinced to ensure a patient’s condition warrants an indwelling catheter, and to consider safer alternatives such as a condom catheter and bladder scanner with intermittent straight catheterization.
2. <u>Upon hospital admission, the indwelling catheter often remains in place</u> o The project leader can share the latest data from the medical unit(s), showing how many of the floor’s Foleys started out in the ED, what percentage were for inappropriate indications, and what percentage led to infection.
  - It is helpful to establish clear guidelines for urinary catheter use in the ED and to educate the staff on the appropriate indications for the catheter and how to use aseptic insertion technique in those patients who truly need the catheter.
  - We have found that the most effective approach is for the project manager and/or the nurse champion (ideally in this case the nurse champion is an ED nurse) to spend a part of each day walking through the ED, reminding everyone they see about the intervention, asking a nurse or a physician whether the Foley they are about to insert is really necessary (i.e., whether it meets the appropriateness criteria). It is especially important to identify patients being admitted to the hospital from the ED to reassess if the indwelling urinary catheter is still appropriate.
3. For more information on urinary catheters and the ED, see “[Appropriate urinary catheter placement in the emergency department“](http://www.catheterout.org/uploads/5/6/5/0/56503399/_ed_urinary_catheter_2012_02.27.2012.pdf.pdf) prepared by our team and funded by the Agency for Healthcare Research and Quality (AHRQ)
4. <u>Further Reading Suggestions</u> o Fakih MG, Heavens M, Grotemeyer J, Szpunar SM, Groves C, Hendrich A. Avoiding potential harm by improving appropriateness of urinary catheter use in 18 emergency departments. Ann Emerg Med.2014;63(6):761-8.
  - Fakih MG, Pena ME, Shemes S, Rey J, Berriel-Cass D, Szpunar SM, Savoy-Moore RT, Saravolatz LD. Effect of establishing guidelines on appropriate urinary catheter placement. Acad Emerg Med.2010;17(3):337–40.
  - Gokula RM, Smith MA, Hickner J. Emergency room staff education and use of a urinary catheter indication sheet improves appropriate use of Foley catheters. AmJ Infect Control.2007;35(9):589-93.

## Appendix B Type the title of your appendix here

JBI Levels of Evidence

*Developed by the Joanna Briggs Institute Levels of Evidence and Grades of Recommendation Working Party October 2013*

PLEASE NOTE: These levels are intended to be used alongside the supporting document outlining their use. Using Levels of Evidence does not preclude the need for careful reading, critical appraisal and clinical reasoning when applying evidence.

### LEVELS OF EVIDENCE FOR EFFECTIVENESS

**Level 1 – Experimental Designs**

Level 1.a – Systematic review of Randomized Controlled Trials (RCTs)

Level 1.b – Systematic review of RCTs and other study designs

Level 1.c – RCT

Level 1.d – Pseudo-RCTs

**Level 2 – Quasi-experimental Designs**

Level 2.a – Systematic review of quasi-experimental studies

Level 2.b – Systematic review of quasi-experimental and other lower study designs Level 2.c –

Quasi-experimental prospectively controlled study

Level 2.d – Pre-test – post-test or historic/retrospective control group study

**Level 3 – Observational – Analytic Designs**

Level 3.a – Systematic review of comparable cohort studies

Level 3.b – Systematic review of comparable cohort and other lower study designs Level 3.c – Cohort study with control group Level 3.d – Case – controlled study

Level 3.e – Observational study without a control group

**Level 4 – Observational –Descriptive Studies** Level 4.a

Systematic review of descriptive studies Level 4.b –

Cross-sectional study

Level 4.c – Case series

Level 4.d – Case study

**Level 5 – Expert Opinion and Bench Research** Level

5.a – Systematic review of expert opinion Level 5.b – Expert consensus

Level 5.c – Bench research/ single expert opinion

### LEVELS OF EVIDENCE FOR DIAGNOSIS

**Level 1 – Studies of Test Accuracy among consecutive patients**

Level 1.a – Systematic review of studies of test accuracy among consecutive patients Level 1.b – Study of test accuracy among consecutive patients

**Level 2 – Studies of Test Accuracy among non-consecutive patients**

Level 2.a – Systematic review of studies of test accuracy among non-consecutive patients Level 2.b – Study of test accuracy among non-consecutive patients

**Level 3 – Diagnostic Case control studies**

Level 3.a – Systematic review of diagnostic case control studies Level 3.b –

Diagnostic case-control study **Level 4 – Diagnostic yield studies**

Level 4.a – Systematic review of diagnostic yield studies Level

4.b – Individual diagnostic yield study

**Level 5 – Expert Opinion and Bench Research** Level

5.a – Systematic review of expert opinion Level 5.b

Expert consensus

Level 5.c – Bench research/ single expert opinion

### LEVELS OF EVIDENCE FOR PROGNOSIS

**Level 1 – Inception Cohort Studies**

Level 1.a – Systematic review of inception cohort studies

Level 1.b – Inception cohort study **Level 2 – Studies of All or none**

Level 2.a – Systematic review of all or none studies

Level 2.b – All or none studies **Level 3 – Cohort studies**

Level 3.a – Systematic review of cohort studies (or control arm of RCT) Level 3.b – Cohort study (or control arm of RCT)

**Level 4 – Case series/Case Controlled/ Historically Controlled studies**

Level 4.a – Systematic review of Case series/Case Controlled/ Historically Controlled studies Level 4.b – Individual Case series/Case Controlled/ Historically Controlled study

**Level 5 – Expert Opinion and Bench Research** Level

5.a– Systematic review of expert opinion Level 5.b

Expert consensus

Level 5.c – Bench research/ single expert opinion

### LEVELS OF EVIDENCE FOR ECONOMIC EVALUATIONS

**Levels**

1. Decision model with assumptions and variables informed by systematic review and tailored to fit the decision making context.
2. ystematic review of economic evaluations conducted in a setting similar to the decision makers.
3. Synthesis/review of economic evaluations undertaken in a setting similar to that in which the decision is to be made and which are of high quality (comprehensive and credible measurement of costs and health outcomes, sufficient time period covered, discounting, and sensitivity testing).
4. Economic evaluation of high quality (comprehensive and credible measurement of costs and health outcomes, sufficient time period covered, discounting and sensitivity testing) and conducted in setting similar to the decision making context.
5. Synthesis / review of economic evaluations of moderate and/or poor quality (insufficient coverage of costs and health effects, no discounting, no sensitivity testing, time period covered insufficient).
6. Single economic evaluation of moderate or poor quality (see directly above level 5 description of studies).
7. Expert opinion on incremental cost effectives of intervention and comparator.

### LEVELS OF EVIDENCE FOR MEANINGFULNESS

1. Qualitative or mixed-methods systematic review
2. Qualitative or mixed-methods synthesis
3. Single qualitative study
4. Systematic review of expert opinion
5. Expert opinion

## Appendix C

**Targeted Assessment for Prevention (TAP) Facility Assessment Tool**

**<u>Catheter-associated Urinary Tract Infection (CAUTI) Targeted Assessment for Prevention (TAP) Facility Assessment Tool</u>**

**Notes for the Respondent:**

- This assessment is meant to capture your *awareness and perceptions of policies and practices* related to catheter-associated urinary tract infection (CAUTI) prevention at the facility or unit in which this assessment is being administered.
- Responses should refer to what is *currently* in place at the facility or unit in which the assessment is being administered.
- Please use the comment boxes to elaborate and capture information as needed - such detailed comments may help focus additional drill down opportunities and next steps.

**Instructions for Submission:**

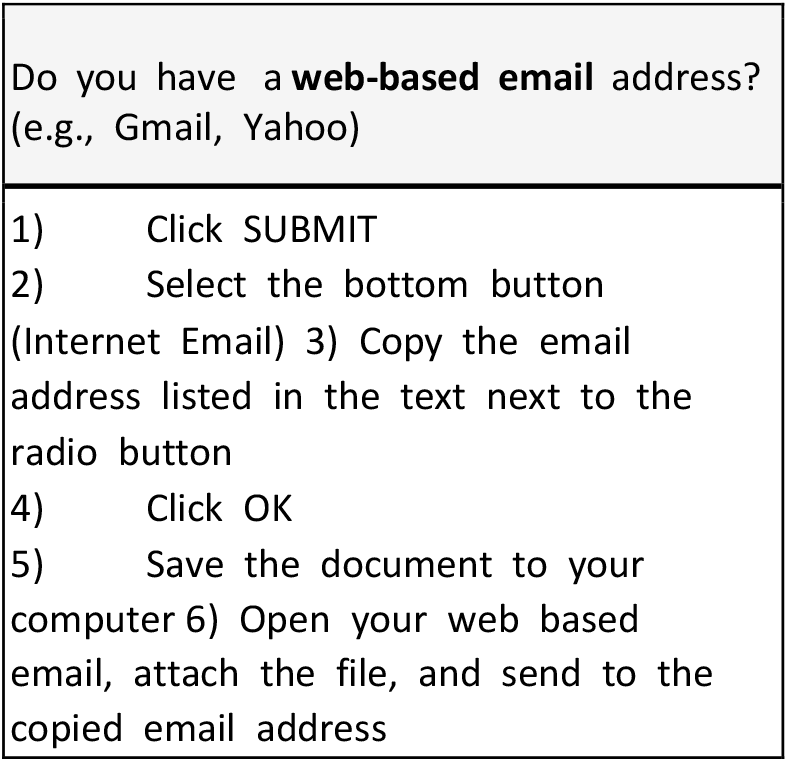

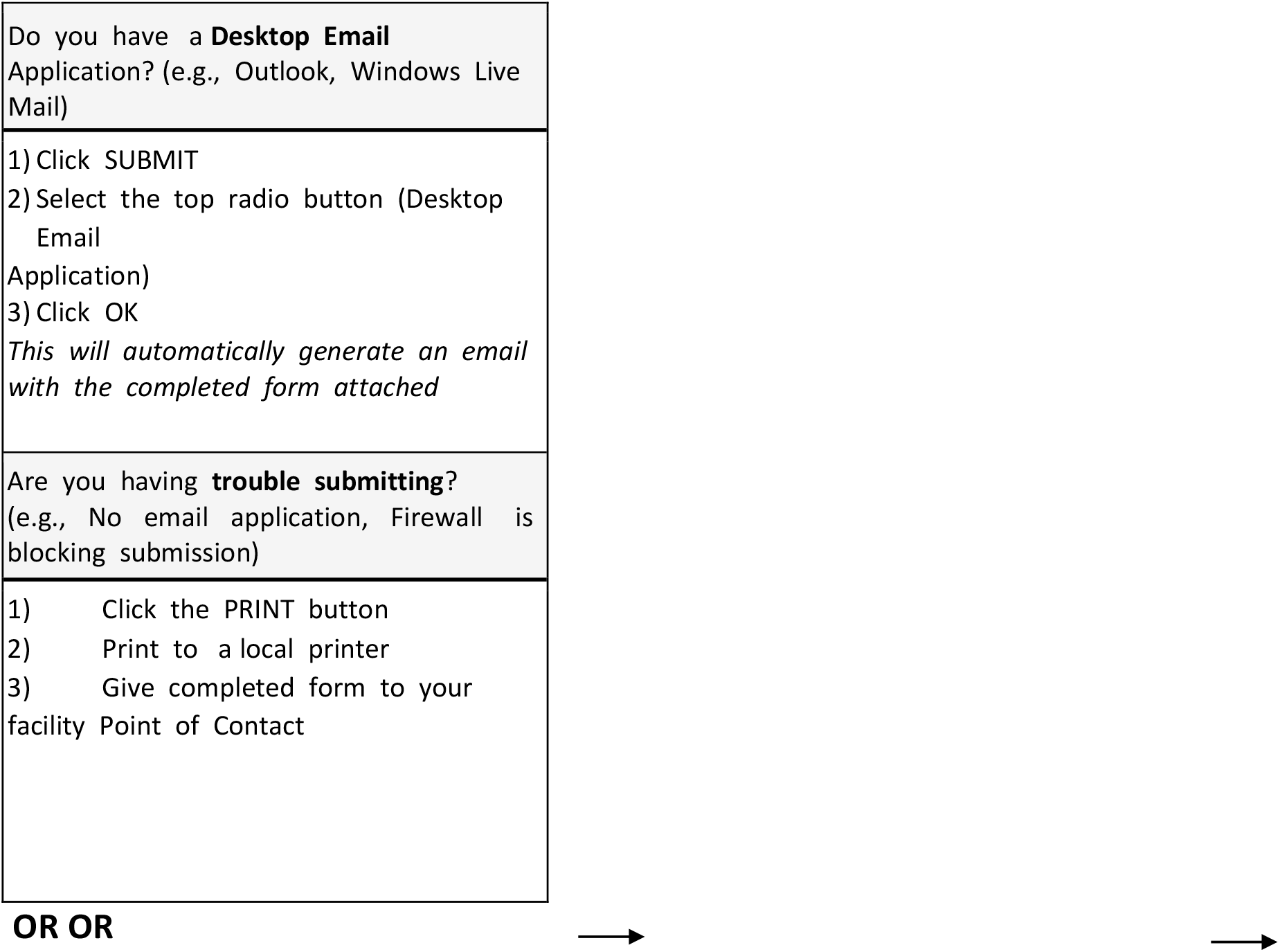

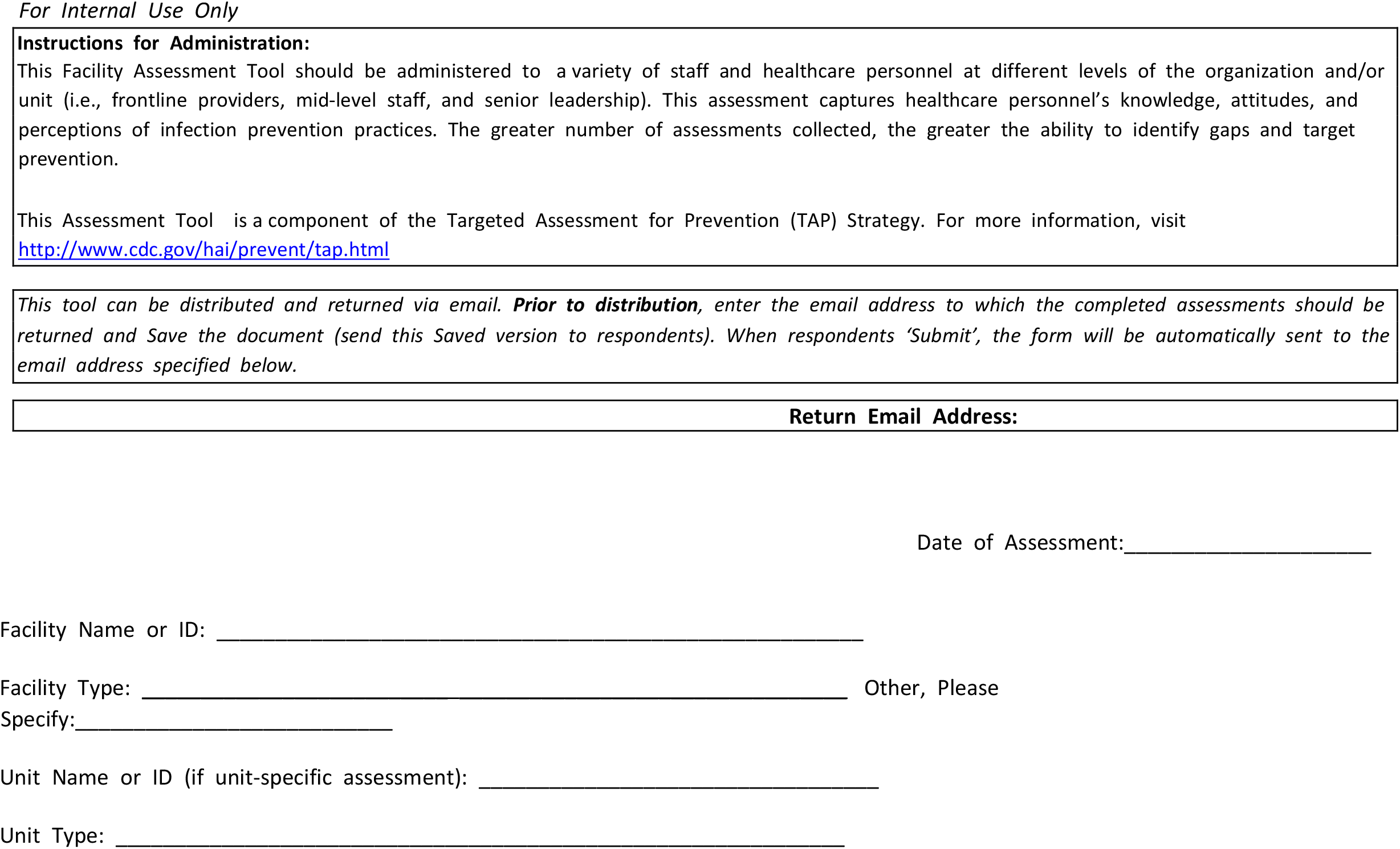

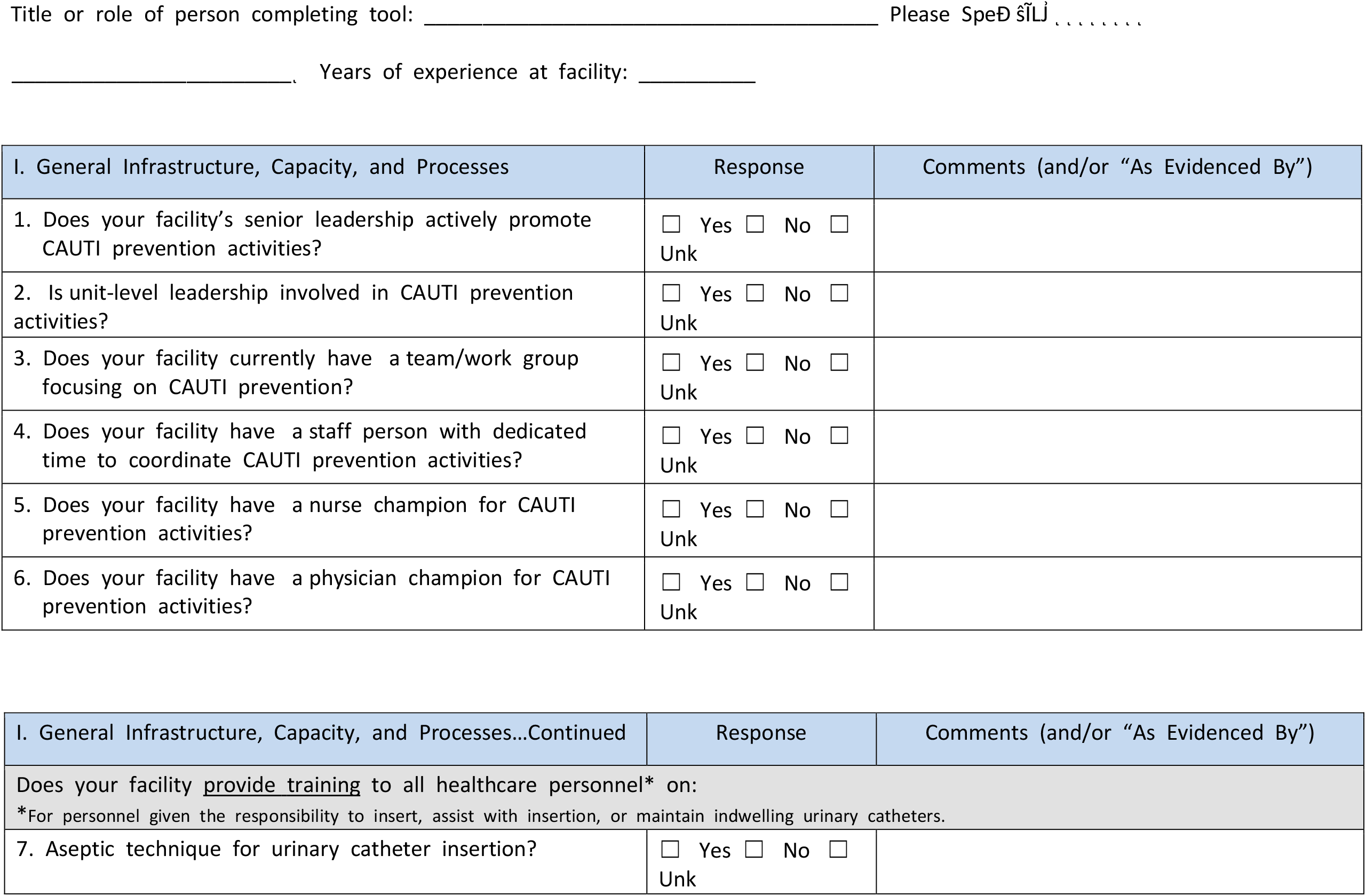
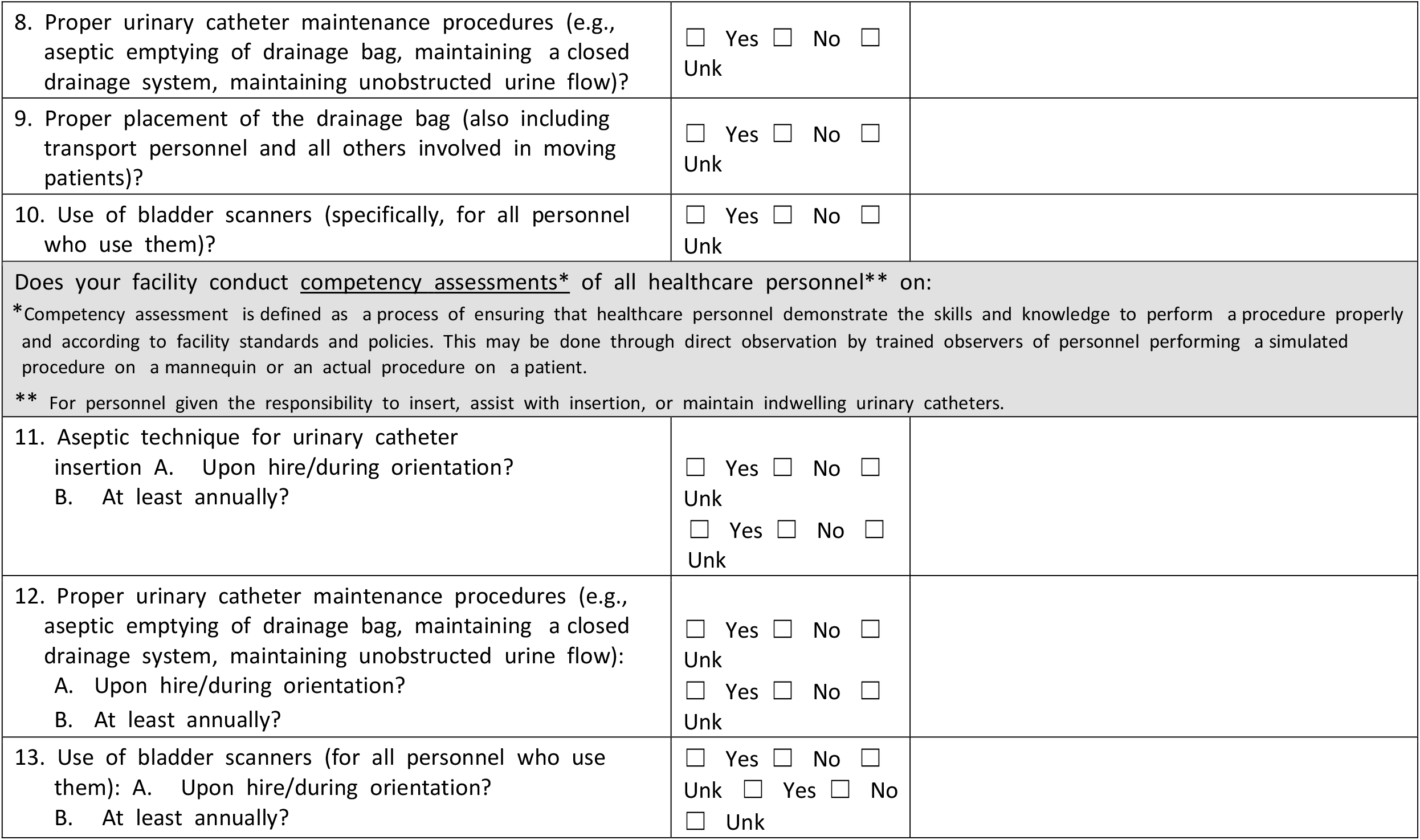

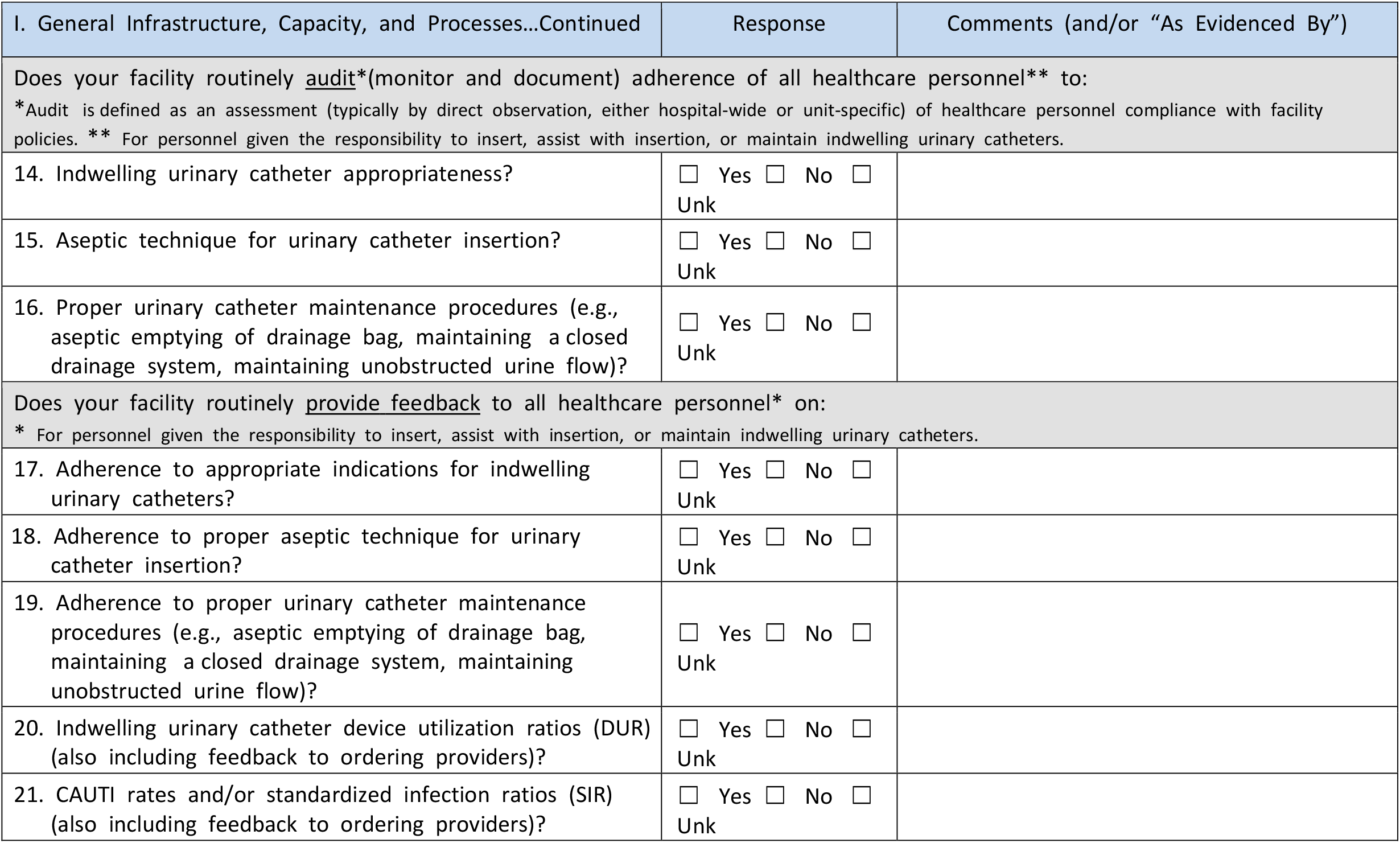

| II. Appropriate Indications for Indwelling Urinary Catheter Insertion | Response Choices |  |  |  |  |  | Comments (and/or "As Evidenced By") |
| --- | --- | --- | --- | --- | --- | --- | --- |
|  |  |  | : |  |  | : |  |
| 1. Do ordering providers document an indication for indwelling urinary catheters? |  |  |  |  |  |  |  |
| 2. Do ordering providers use indwelling urinary catheters for appropriate indications? |  |  |  |  |  |  |  |
| 3. Do personnel use alternative strategies for management of urinary incontinence (e.g., external catheters, bedside commodes, scheduled toileting, garments/pads)? |  |  |  |  |  |  |  |
| 4. Do personnel use bladder scanners to confirm urinary retention before placing or replacing urinary catheters? |  |  |  |  |  |  |  |
| 5. Do personnel use bladder scanners with intermittent catheterization for management of postoperative urinary retention? |  |  |  |  |  |  |  |
| 6. Does your facility provide instructions/protocols for personnel to act upon bladder scanner results? |  |  |  |  |  |  |  |
| 7. Are patients and/or families educated on appropriate indications for and care of urinary catheters? |  |  |  |  |  |  |  |
| 8. In the Emergency Department, is an order provided prior to insertion of an indwelling urinary catheter? |  |  |  |  |  |  |  |
| 9. Do Emergency Department providers order indwelling urinary catheters for appropriate indications? |  |  |  |  |  |  |  |
| 10. Do Emergency Department providers document an indication when ordering indwelling urinary catheters? |  |  |  |  |  |  |  |

| III. Aseptic Indwelling Urinary Catheter Insertion | Response Choices |  |  |  |  |  | Comments (and/or "As Evidenced By") |
| --- | --- | --- | --- | --- | --- | --- | --- |
|  |  |  | : |  |  | : |  |
| 1. Are supplies/kits for proper aseptic indwelling urinary catheter insertion available in all patient care locations where urinary catheters are inserted? |  |  |  |  |  |  |  |
| 2. Does your facility require at least two personnel to be present for indwelling urinary catheter insertions – one to perform the insertion and the other(s) to observe the procedure to ensure proper aseptic technique (e.g., using a checklist)? |  |  |  |  |  |  |  |
| 3. Does the person inserting the indwelling urinary catheter document the insertion procedure (e.g., date, person[s] performing procedure, complications)? |  |  |  |  |  |  |  |

| IV. Proper Indwelling Urinary Catheter Maintenance | Response Choices |  |  |  |  |  | Comments (and/or "As Evidenced By") |
| --- | --- | --- | --- | --- | --- | --- | --- |
|  |  |  | : |  |  | - |  |
| 1. Does your facility utilize pre-connected, sealed urinary catheter drainage systems? |  |  |  |  |  |  |  |
| 2. Do personnel keep the urinary drainage system closed to maintain sterility (e.g., do not perform open irrigation of the urinary catheter)? |  |  |  |  |  |  |  |
| 3. In the Emergency Department, are pre-connected, sealed urinary drainage systems <u>with urine meters</u> used in critically ill patients to avoid breaking the system once transferred to the intensive care unit? |  |  |  |  |  |  |  |

| V. Timely Removal of Indwelling Urinary Catheters | Response Choices |  |  |  |  |  | Comments (and/or "As Evidenced By") |
| --- | --- | --- | --- | --- | --- | --- | --- |
|  |  |  | : |  |  | - |  |
| 1. Does your facility identify patients who have indwelling urinary catheters in place (e.g., in the electronic medical record or a daily unit list)? |  |  |  |  |  |  |  |
| 2. Are patients with indwelling urinary catheters reviewed daily for continued need (e.g., by bedside nurses and/or interdisciplinary team)? |  |  |  |  |  |  |  |
| 3. Are indwelling urinary catheters removed in the postanesthesia care unit (PACU) if there is no indication for continued use after surgery? |  |  |  |  |  |  |  |
| 4. Does your facility use alerts, reminders, or stop orders for indwelling urinary catheter removal? |  |  |  |  |  |  |  |
| 5. *If applicable*, do physicians respond to alerts or reminders by removing unnecessary urinary catheters? |  |  |  |  |  |  | *Not Applicable* |
| 6. *If applicable*, do nurses respond to alerts or reminders by removing unnecessary urinary catheters or calling the physician? |  |  |  |  |  |  | *Not Applicable* |
| 7. If a nurse-directed removal protocol is in place, do nurses remove indwelling urinary catheters at your facility? |  |  |  |  |  |  | *Not Applicable* |
| 8. If a nurse-directed removal protocol is in place, are nurses comfortable using it? |  |  |  |  |  |  | *Not Applicable* |
| 9. If a nurse-directed removal protocol is in place, are physicians supportive of nurses using it? |  |  |  |  |  |  | *Not Applicable* |

| VI. Appropriate Urine Culturing Practices | Response Choices |  |  |  |  |  | Comments (and/or "As Evidenced By") |
| --- | --- | --- | --- | --- | --- | --- | --- |
|  |  |  | : |  |  | - |  |
|  |  |  | : |  |  | - |  |
| 1. Do ordering providers send urine cultures only on patients with signs or symptoms of a urinary tract infection? |  |  |  |  |  |  |  |
| 2. If a urine culture is ordered, is a urinalysis ordered at the same time? |  |  |  |  |  |  |  |
| 3. Are urine culture specimens transported to the lab for testing immediately following collection (i.e., not allowed to sit unrefrigerated for prolonged periods of time)? |  |  |  |  |  |  |  |

Additional Comments/Observations (Please specify/describe in detail):

## Appendix D

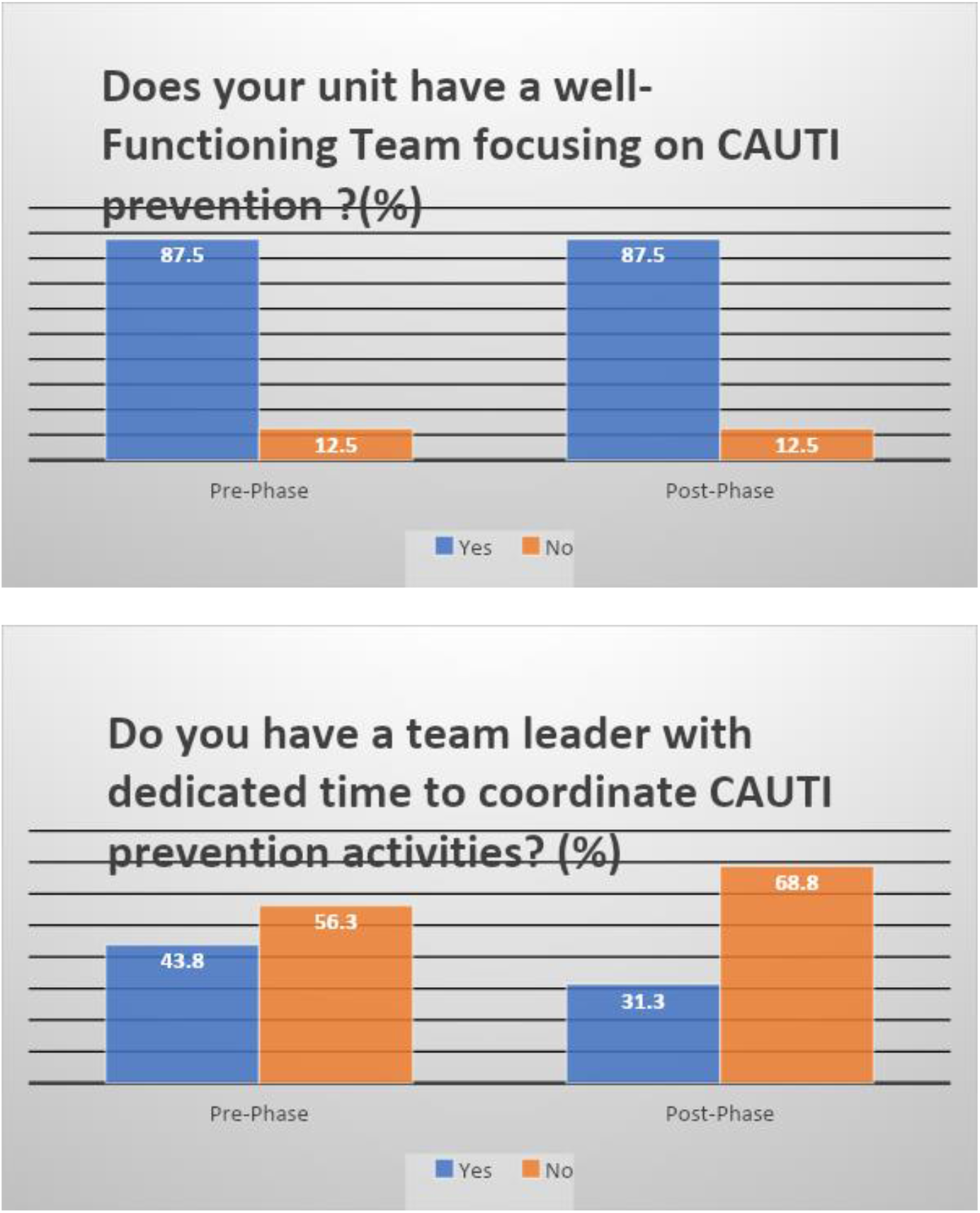

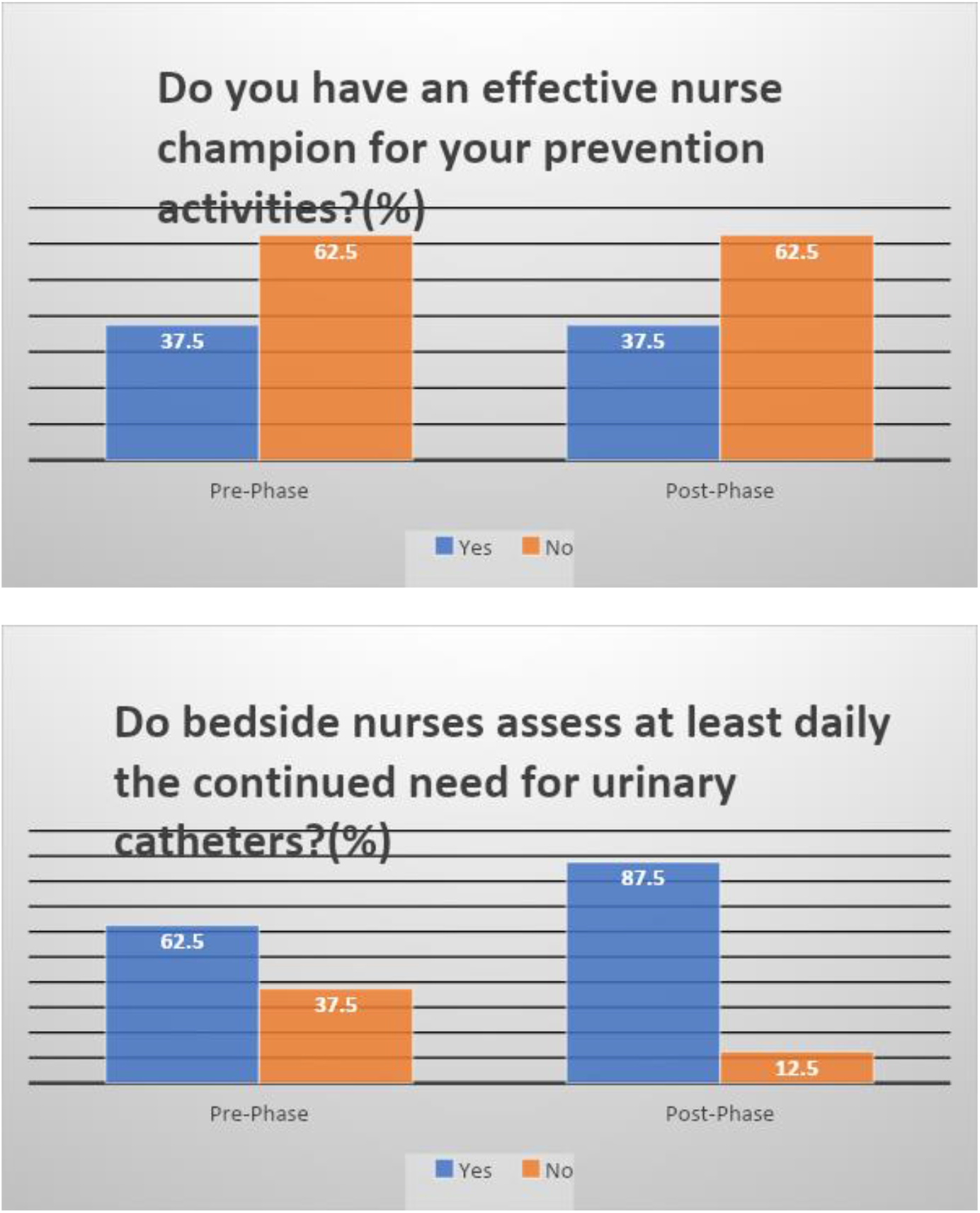

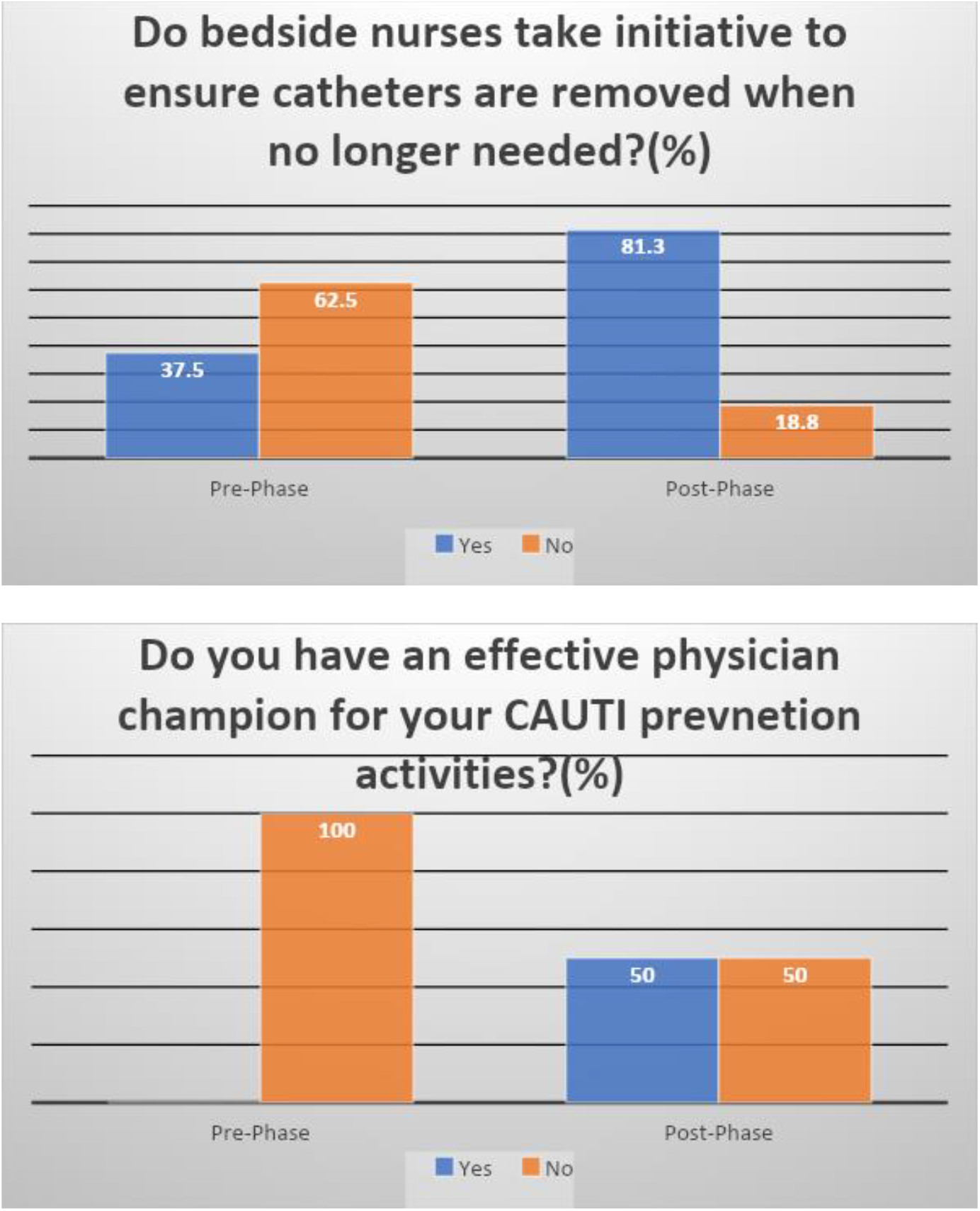

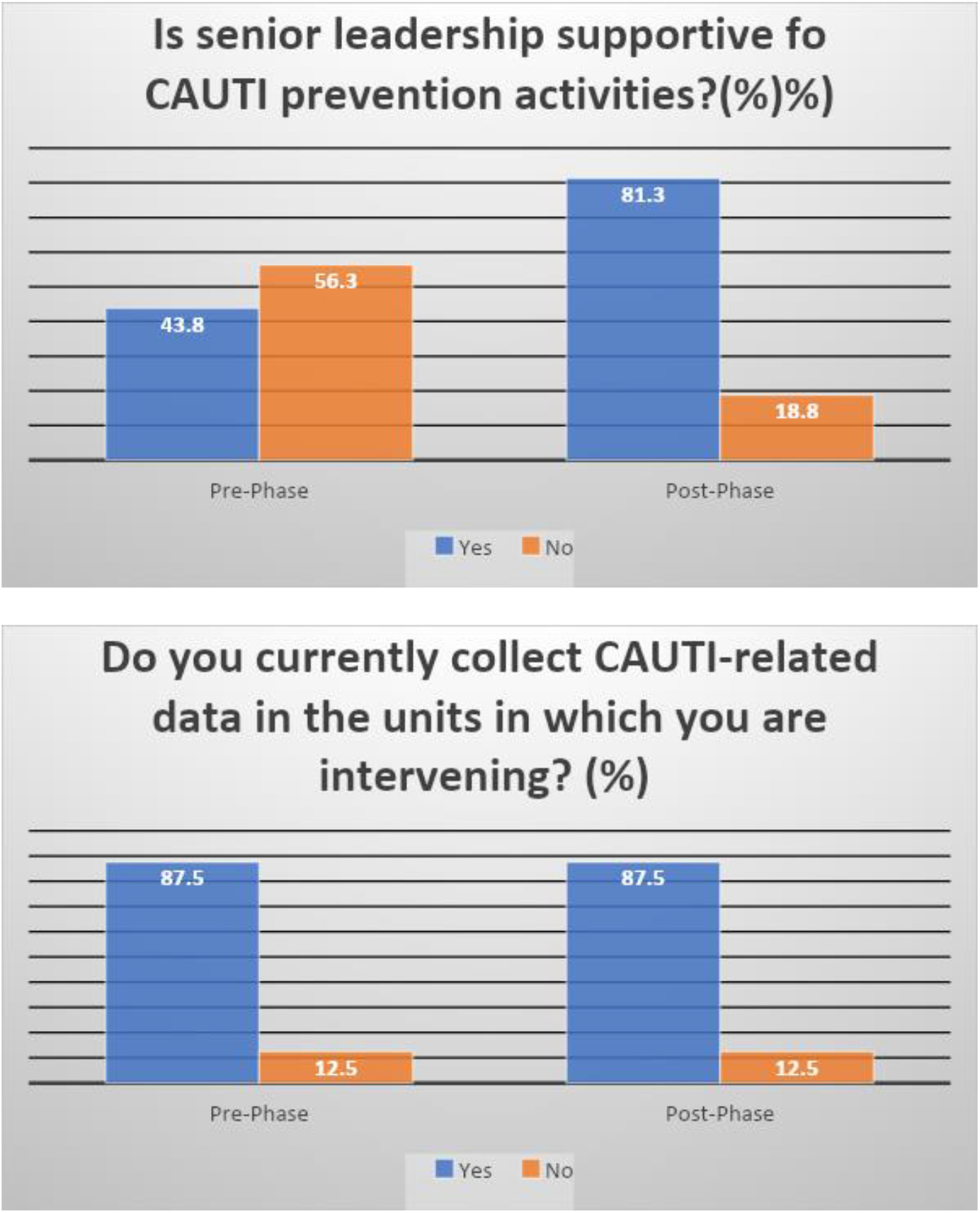

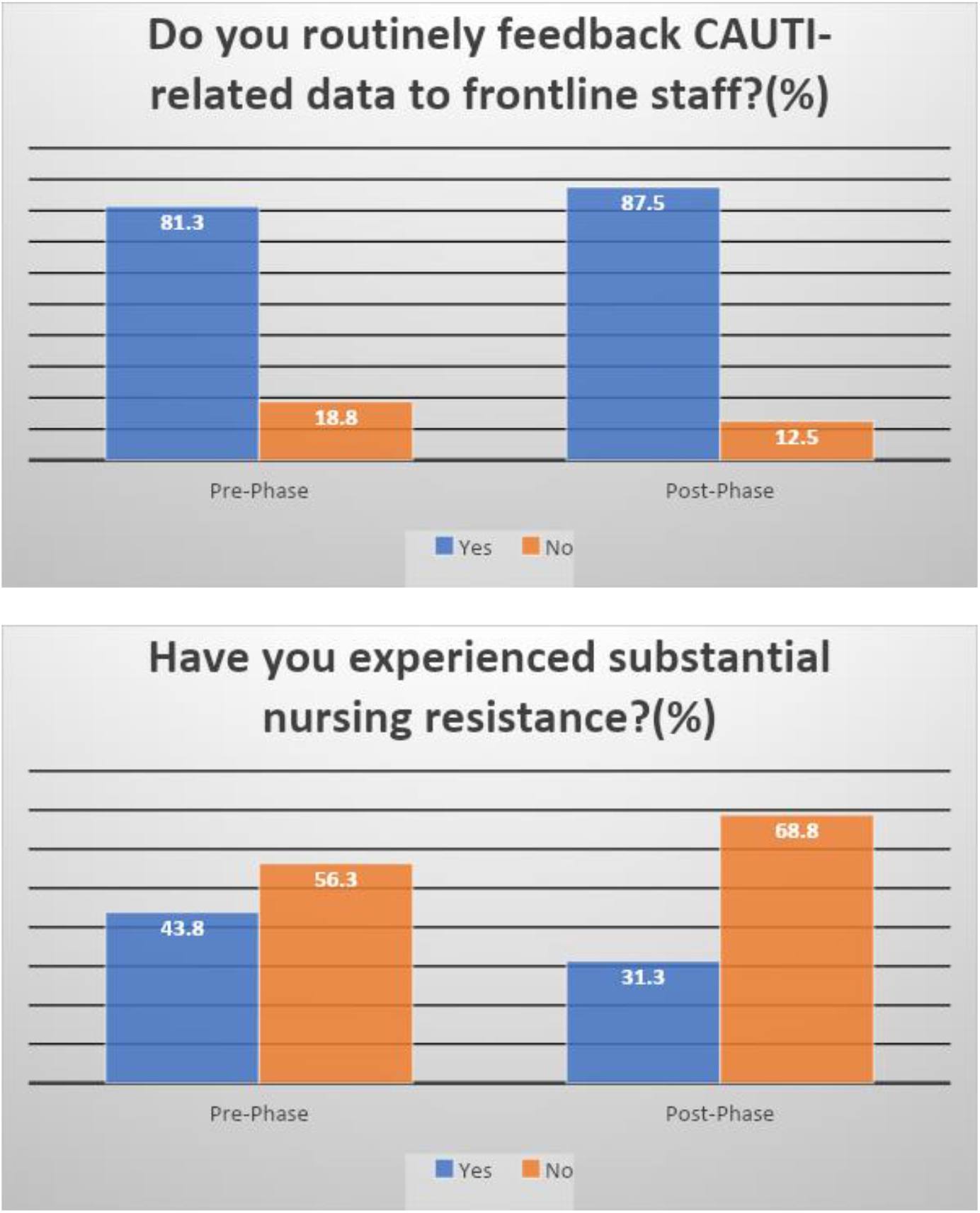

